# The path toward generalizable clinical prediction models

**DOI:** 10.1101/2024.04.16.24305902

**Authors:** Fredrik Hieronymus, Magnus Hieronymus, Axel Sjöstedt, Staffan Nilsson, Jakob Näslund, Alexander Lisinski, Søren Dinesen Østergaard

## Abstract

The peaking phenomenon refers to the observation that, after a point, the performance of prediction models starts to decrease as the number of predictors (p) increases. This issue is commonly encountered in small datasets (colloquially known as “small n, large p” datasets or high-dimensional data). It was recently reported based on analysis of data from five placebo-controlled trials that clinical prediction models in schizophrenia showed poor performance (average balanced accuracy, BAC, 0.54). This was interpreted to suggest that prediction models in schizophrenia have poor generalizability. In this paper we demonstrate that this outcome more likely reflects the peaking phenomenon in a small n, large p dataset (n=1513 participants, p=217) and generalize this to a set of illustrative cases using simulated data. We then demonstrate that an ensemble of supervised learning models trained using more data (18 placebo-controlled trials, n=4634 participants), but fewer predictors (p=33), achieves better prediction (average BAC = 0.64) which generalizes to out-of-sample studies as well as to data from active-controlled trials (n=1463, average BAC = 0.67). Based on these findings, we argue that the achievable prediction accuracy for treatment response in schizophrenia— and likely for many other medical conditions—is highly dependent on sample size and the number of included predictors, and, hence, remains unknown until more data has been analyzed. Finally, we provide recommendations for how researchers and data holders might work to improve future data analysis efforts in clinical prediction.

---

Being able to tell a patient prior to treatment initiation that while two different treatments are on average equally efficacious, *they* will likely respond favorably to just one of them whereas the other would probably mainly result in side effects, is the holy grail of precision medicine. ^1^ While individualized treatment based on not only tumor, but also patient characteristics is becoming a reality for a number of cancers^2,3^ there are few parallels in other fields of medicine, and certainly none for any major psychiatric disorder. ^4^ For psychiatry, it is doubtful that any strong and simple (here meaning linear and additive) predictors remain to be identified in currently available data. If there were such relations to, e.g., commonly studied blood markers, sociodemographic variables, symptomatology, genetic information, etc., then they would likely already have been discovered since not much has changed in terms of how we delineate or treat major psychiatric disorders over the last decades. ^5^ Naturally, then, focus has shifted toward exploring weak and/or complex (i.e., non-linear and/or non-additive) predictors, and, specifically, to using machine learning methodology without pre-specifying predictor-outcome relations.

Supervised learning models are a subset of machine learning models which have been commonly employed for clinical prediction in psychiatry. ^6^ In supervised learning, the outcome of interest—e.g., will this patient exhibit a clinically relevant outcome after four weeks of treatment—is known and the models are tasked with approximating the function that best maps the input data (predictors) to the outcome. Such models, given reasonable parameters regarding, e.g., cross-validation strategy and hyperparameter tuning, tend to perform very well when the number of study subjects (n) is much higher than the number of predictors (p)—colloquially known as “large n, small p” datasets—but less well when there is a large number of predictors relative to the number of subjects (small n, large p). ^7,8^

Chekroud and co-workers recently reported that supervised learning models (elastic net, p=217; random forest, p=137) trained to predict symptom remission in schizophrenia using training data from four randomized controlled trials (n=1032 to 1414 participants; depending on which study was withheld from training) failed to predict symptom remission in a holdout trial at a level better than chance (average balanced accuracy, BAC: 0.54 for elastic net; 0.53 for random forest). ^9^ From this, Chekroud et al. concluded that models predicting treatment outcomes in schizophrenia are highly context-dependent and have limited generalizability between trials. We believe that this conclusion is too pessimistic and instead reflects the peaking phenomenon, i.e., that after a point (peak), the performance of prediction models decreases as the number of predictors increases^7,8,10^

Using a three-times larger schizophrenia dataset (18 trials, n=4634 participants) from the same data sharing organization – Yale University Open Data Access project, YODA^11^ – which includes the five studies analyzed by Chekroud and co-workers, we demonstrate that their reported classification accuracy (average BAC: 0.54/0.53) is compatible with subsamples randomly drawn from the larger case population and thus not contingent on between-study differences. We further illustrate, using actual and simulated data, that the limited performance is likely due to the peaking phenomenon in that neither elastic net nor random forest models perform well in the presence of a) uninformative noisy predictors and b) weak predictors. We end by demonstrating that, using the larger schizophrenia dataset referred to above and fewer predictors (p=33), it is possible to construct an ensemble of supervised learning models that achieves better prediction (average BAC = 0.64) that generalizes to out-of-sample studies, as well as to data from active-controlled antipsychotic trials.

## Data sources

We requested patient-level data for all industry-sponsored, acute-phase, placebo-controlled trials of risperidone and/or paliperidone via the YODA project. ^11^ Remote access to patient-level data was provided by Johnson & Johnson and YODA for all 19 requested studies. One study (RIS-USA-1/Study 201) did not use the Positive and Negative Syndrome Scale (PANSS) ^12^ for symptom rating (which was used both as predictors and outcome) and could hence not be included. The remaining 18 trials constitute the train-and-test dataset. We also requested data for three acute-phase trials of risperidone and/or paliperidone that did not include a placebo-group. These three active-controlled trials (n=1463) served as an extender dataset and were used to assess generalizability.

## Analytic approach

All analyses were run on R version 4.3.0 via the YODA Remote Desktop environment. All models were trained using the Classification and Regression Training (caret) package. Models were tasked to maximize the area under the receiver operating curve (AUC-ROC) and used ten-fold cross-validation. Symptom remission, as defined by the Remission in Schizophrenia Working Group, ^13^ after four weeks of treatment was the outcome parameter for all analyses. BAC – the mean of sensitivity and specificity – is reported for all analyses. For more detailed information regarding, e.g., hyperparameter tuning and simulation parameters, please consult the SM.

We first assessed whether the single-study accuracy figures reported by Chekroud and colleagues were contingent on between-study heterogeneity, or if those estimates were compatible with chance. This was done by Monte Carlo subsampling the test-and-train set 50 times (25 times for random forest) into training and test sets equal in size to the five in- and out-sample groupings used by Chekroud et al. For each subsampled training set, each test set was resampled 100 times using non-parametric bootstrapping. Elastic net and random forest models were implemented as specified by Chekroud and co-workers, however these models were trained either a) using a sparse predictor set consisting of the 30 PANSS-rated symptoms at baseline, treatment (antipsychotic/placebo), age and sex (total p=33) or b) using a noisy predictor set which included the sparse predictor set as well as simulated noisy (here meaning uncorrelated to the outcome; see SM for details) predictors. The latter was done to get the same number of predictors (p=217 or 137, respectively) as in the analyses by Chekroud et al. ^9^

To illustrate the dependence on n, we simulated data sets of different sizes (n=2000 to 25000) that included multiple (p=50) random normal predictors that were weakly (r=0.050, 0.100 and 0.150) correlated to a random normal variable (Y) representing the outcome of interest. To replicate a classification problem, Y was then dichotomized into cases (Y > 0) and non-cases (Y < 0). We also simulated a variable number (p=150 or 450) of noisy predictors. The simulated datasets were split (80:20) into training and test sets and elastic net and generalized linear models were trained. Classification performance was compared to the theoretical performance achievable by a model using the sum of all informative predictors as the only classifier (see SM for details).

We then trained an ensemble of five machine learning models: Classification and Regression Training (CART; caret method: treebag), elastic net (caret method: glmnet), generalized linear model (glm) (caret method: glm), random forest (caret method: rf) and extreme gradient boosting trees (caret method: xgbTree) using the full train-and-test set. To assess the dependence on n, ensemble models were trained using Monte Carlo subsampled training sets varying in size between 384 and 4384 cases, with 50 subsamples per training set size. Model performance was tested against a) 100 subsampled or bootstrapped test sets (n=250) and *b)* 100 bootstrapped extender data sets (n=1463). We also performed single-study testing in which 17 trials were used for training and the test set was the single-study not used for training.

## Description of dataset

In brief, the train-and-test dataset consists of 18 placebo-controlled trials in which 6689 participants were randomized and where 4634 individuals (n range=98 to 471 per trial) had outcome data available at week four. The four-week retention rate was 61% for those randomized to placebo (1097 of 1796) and 72.2% for those randomized to antipsychotics (3537 of 4893). For those with outcome data, symptom remission after four weeks was attained by 36.4% of those receiving antipsychotics (1286) and by 31.1% of those receiving placebo (341). The extender set consists of 3 active-controlled trials in which 1725 participants were randomized and where 1463 individuals (84.8%) had outcome data available after six weeks of treatment. Symptom remission was attained by 42.6% (623) of those with available outcome data. More thorough descriptions of all included studies, as well as of relevant patient characteristics, can be found in the SM.

### Contrasting single study and random subsample response prediction estimates

Figure 1 shows the single-study response prediction estimates reported by Chekroud et al. and correspondingly sized Monte Carlo subsample response prediction estimates. All single-study estimates for the elastic net model lie within the 95% confidence intervals (CIs) for both the sparse and noisy predictor sets. For the random forest model, all single-study estimates lie within the 95% CIs for the noisy predictor set while two out of five had worse than expected BAC compared to the sparse predictor set. Prediction accuracy was significantly decreased by the inclusion of noisy predictors across all training sizes, and more so for random forest than elastic net. CIs increased with training set size reflecting increased variability with decreasing test set size.

**Figure 1.**
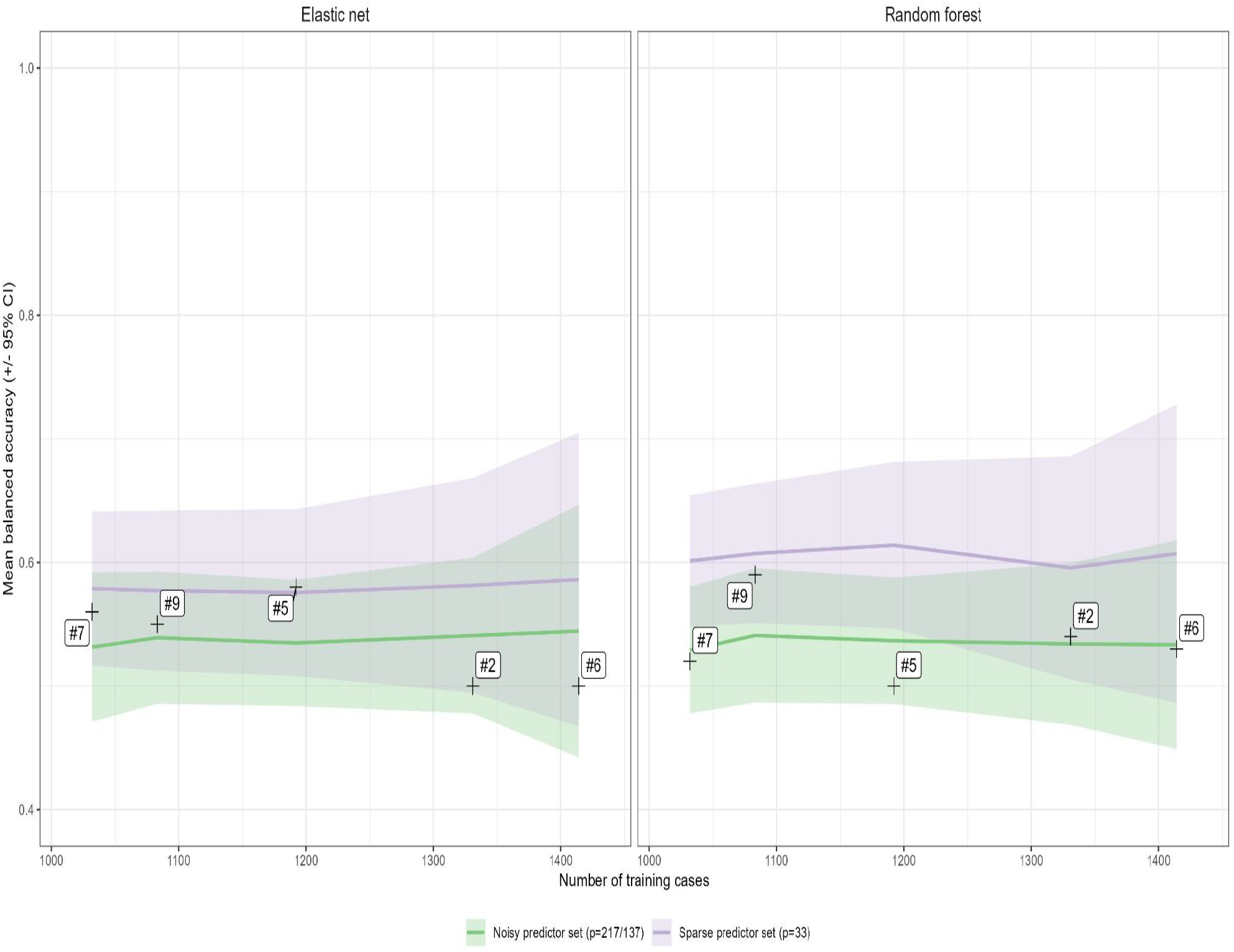
Four-week remission prediction: comparison between subsampled and single-study accuracy for noisy and sparse predictor sets. Solid lines give mean subsampled balanced accuracy and ribbons give 95% CIs for 50 Monte Carlo subsamples (25 for random forest) of the same train and test set sizes as used by Chekroud et al. Test sets were bootstrapped 100 times for each subsampled training set. Each number (#2, #5, #6, #7 and #9) represents one of ten single-study accuracies reported by Chekroud et al. in Supplementary tables 5 and 13. ^9^ For a mapping of our reference numbers to the study designations used by Chekroud et al., please consult the SM.

### Feature selection performance of elastic net in simulated data with weak predictors

Figure 2 shows the performance of elastic net and generalized linear model for a selection of predictor-outcome correlations and noisy data, as n increases. With the average correlation of informative predictors set to 0.15, the elastic net model performs much better than the generalized linear model. When 1600 cases are used for training, elastic net achieved 94% of expected accuracy in the presence of 75% noisy predictors (glm: 85%) and 93% with 90% noisy predictors (glm: 66%). With the average correlation set to 0.05, elastic net achieved 57% of expected accuracy in the presence of 75% noisy predictors (glm: 57%) and 41% with 90% noisy predictors (glm: 31%).

**Figure 2.**
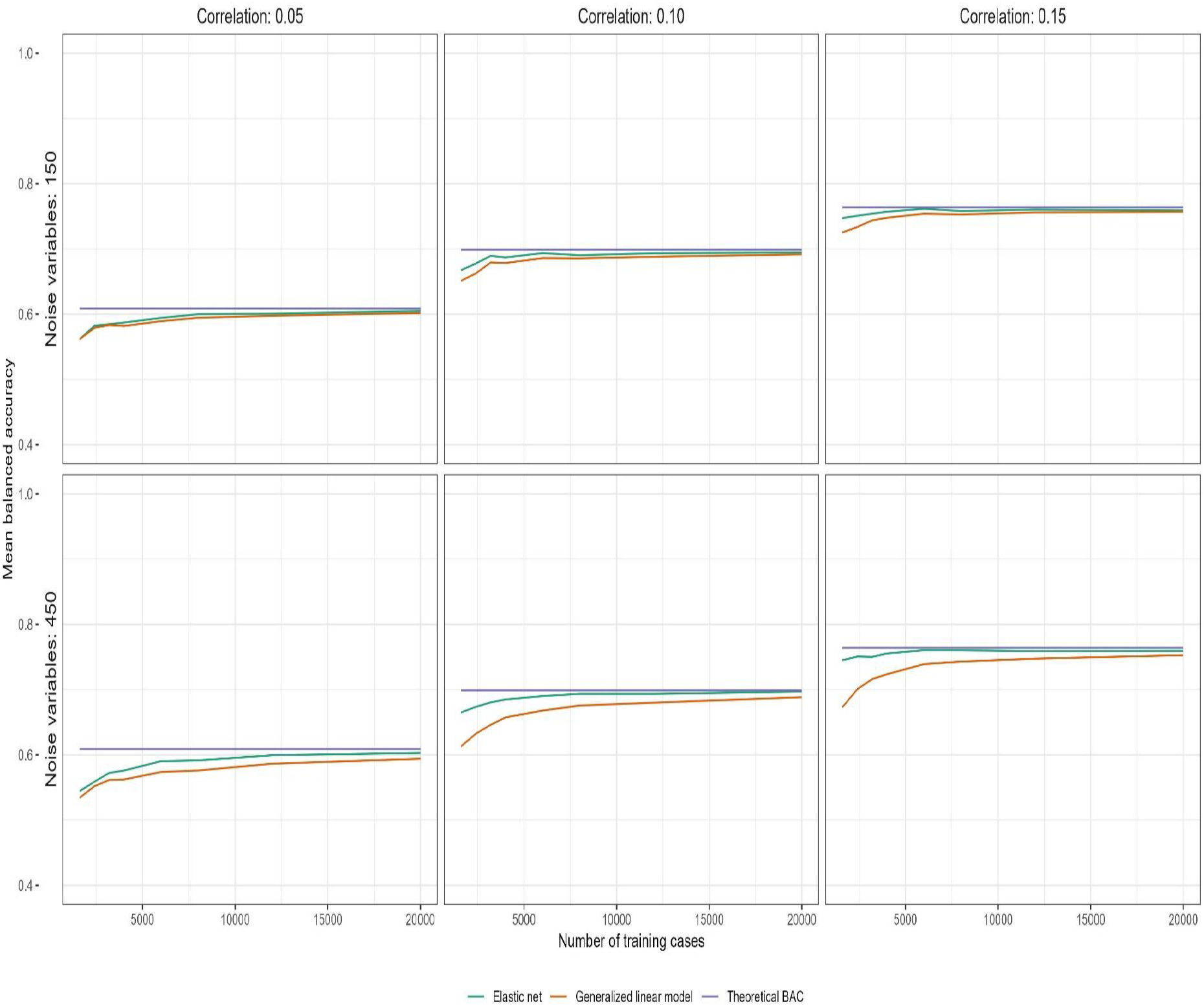
Performance of elastic net and generalized linear models in simulated data with weak predictors. The purple lines shows theoretical BAC for the sum of all informative predictors (p=50). The orange lines show test-set BAC achieved by a generalized linear model and the green lines show the same for elastic net (means are averaged over 50 simulations per training set size).

### Ensemble prediction

Ensemble model BAC for Monte Carlo subsampled test-sets increased from 0.60 to 0.64 as the study population used for training increased from 384 to 4384. For the extender set, BAC correspondingly increased from 0.63 to 0.67 (Figure 3). The performance increase in the extender set was mainly due to the models better predicting true positives (see SM). The 95% CIs did not overlap with 0.50 (i.e., no difference) at any point. Sixteen out of eighteen leave-one-study-out estimates were within the 95% CIs for Monte Carlo subsampled sets of corresponding sizes. Extreme gradient boosting was the best performing individual model and elastic net the worst. Additional metrics and results for all individual models are found in the SM.

**Figure 3.**
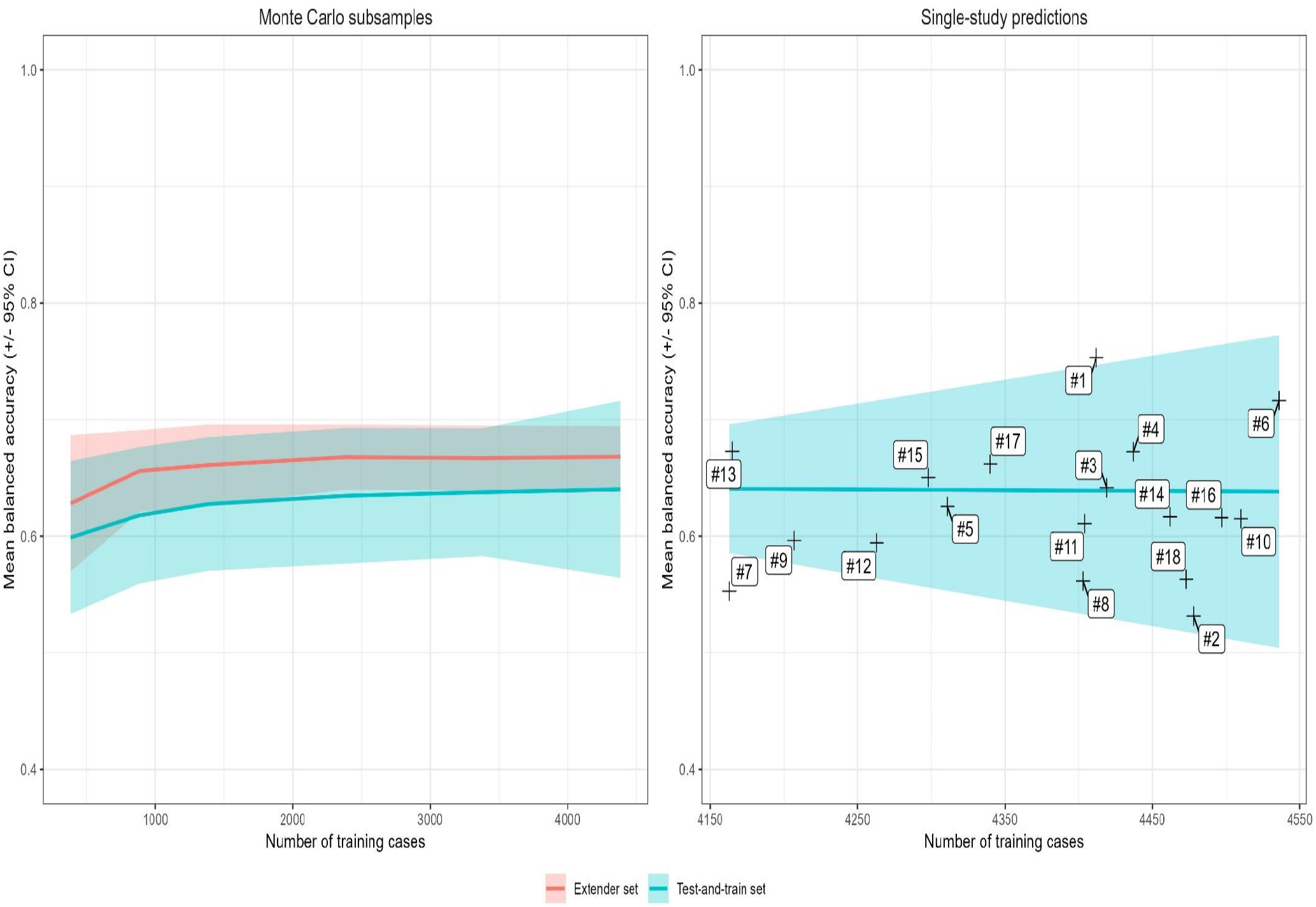
Four-week remission prediction: BAC for ensemble predictions of subsampled test data, extender test data, and single-study data. The left panel gives mean and 95% CI BAC for subsampled training sets ranging in size between 384 to 4384 cases. The right panel shows the same for subsampled and single-study BAC for leave-one-study-out analyses. Each number #1 to #18 represents one study in the train-and-test set (see SM for mapping).

### Interpretation

A supervised learning model trained using a small set of schizophrenia trial data can yield response prediction that outperforms chance, and which generalizes to out-of-sample trials and other trial contexts (active-controlled trials). There was a non-trivial amount of “complex” features (non-linear and/or non-additive) present in the data as evidenced by the greater performance of cart, extreme gradient boosting and random forest over elastic net and glm, respectively. The steep decline in prediction accuracy when including simulated noisy predictors—especially in the presence of very weak predictors (r=0.05)—illustrates that model parsimony and predictor selection are highly important when designing and interpreting supervised learning models. In the cases studied here, relying on the embedded feature selection of the elastic net algorithm was not sufficient to avoid significant performance losses. ^14^ While increasing n solves most issues, this is usually not possible for individual researchers and different strategies for lowering p should routinely be included as sensitivity analyses.

### Future directions for researchers

That better classification performance can be achieved using fewer predictors (peaking phenomenon) is well-known. ^7,8,10,15^ The present result do, therefore, not imply that the predictors included by Chekroud and colleagues, but not by us, are uninformative. Hence, there are many potential predictors left to evaluate and researchers limited by small n datasets should consider exploring such feature spaces systematically—favoring parsimony— rather than using an “all at once” approach that relies solely on models with embedded feature selection. While, e.g., the elastic net algorithm is considered especially useful in small n, large p datasets, ^14^ it did not perform well here.

Even if, as in our noisy predictor set, it is known that 184 out of 217 potential predictors are uninformative, the number of possible ways of selecting 33 variables out of 217 exceeds 10^39^. These selection problems can hence not be brute forced. While feature selection methods^16^ should routinely be assessed when working with small n, large p data sets – our results are a reminder of the value of hypothesis-driven research and domain knowledge also in data-driven research. ^17^ It is evident from previous research that baseline symptomatology is likely to be of importance for outcome prediction. ^18^ It is less evident that, e.g., a panel of standard blood tests hold the same promise. Including reasonably sized sub-analyses of qualitatively different predictor subsets is a hypothesis-driven way in which some of the problems with small n, large p datasets might be mitigated. Predictor subsets showing promise could then be subjected to feature selection and/or dimensionality reduction techniques, and subsequently aggregated into pooled analyses making such analyses less costly in terms of processing power. Naturally, such partitioning runs the risk of missing predictor interactions, but—as here demonstrated—so does the “all at once”-approach and there is nothing stopping both approaches from being employed.

### Future directions for data sharing organizations and data holders

Our analyses are far from exhaustive and the upper limit on the prediction accuracy that might be achieved using the data available for pooling is unknown. While independent researchers can remove a fair bit of this uncertainty by conducting more comprehensive analyses, there are some barriers that only data sharing organizations and data holders can remove, namely the disconnected data landscape and limitations of the research environments.

Our analyses include data from the two antipsychotic development programs (risperidone and paliperidone) that are available via YODA. Thanks to the collaboration between YODA and Vivli.org^19^ it would now be possible to include data also from the development programs for aripiprazole, lurasidone, and olanzapine. While an improvement, many data holders (e.g., Sanofi, amisulpride; AstraZeneca, quetiapine; Pfizer, ziprasidone; Lundbeck, brexpiprazole; AbbVie, cariprazine; Bristol-Myers Squibb, lumateperone; Merck Sharpe & Dohme, asenapine; Vanda, iloperidone) have still either not made the relevant data available or have not made them available in a form that allows for pooling. Given the critical dependence on n for model performance, a better integrated data landscape would be much preferable.

The Remote Desktop environment provided by YODA has some limitations. We were unable to implement parallel processing in R, thus significantly increasing the duration of all analyses and limiting the number and depth of models deemed feasible to explore. We were, e.g., unable to assess Extreme Gradient Boosting with Dropouts meet Multiple Additive Regression Trees (DART; caret package: xgbDART) ^20^ due to a lack of processing power and were unable to implement neural network analyses using keras in R due to software restrictions. The research environment occasionally suffered from severe latency and scheduled maintenance at times collided with ongoing runs which could lead to the loss of days of analysis work.

Some of our issues could have been mitigated by better coding practices and/or by communication with YODA staff (who, it should be noted, have been extremely helpful). However, these are not issues we would have had if the data were available for offline use. While there are privacy and data safety concerns, ^21^ some data sharing organizations— including the National Institutes of Health^22^—already allow for offline data use. Considering that analyses are bound to get larger and more computationally intensive, establishing and adopting procedures for offline data use is bound to greatly improve future research efforts.

An alternative option is for the data sharing organizations to further improve platform interoperability. If done in tandem with allowing researchers to purchase larger allocations of computer resources (as Vivli.org has done), that would also solve many of the issues currently preventing more informative analyses from being done.

### Future analyses

There are several ways in which predictive accuracy might be further improved. The obvious first step is to conduct analyses on larger data sets, including more predictors and using more complex model architecture. Similarly, including data on early treatment response (e.g., after one week of treatment) is bound to improve accuracy and may be nearly as clinically useful (e.g., for getting early guidance on treatment switching). ^23,24^ Training models to predict outcomes other than very good treatment responses, e.g., treatment attrition and adverse events, is also of interest. Such outcomes are relevant by themselves, but they are also not necessarily best predicted by the same features that predict very good efficacy. Hence, coupling models trained to predict very poor, or for that matter very average outcomes, to models trained to predict very good outcomes might yield overall increases in prediction accuracy. ^25^ A separate but important question is what makes these models work, i.e., which features that allow them to be predictive. Reverse engineering the models to, e.g., gauge the extent to which they are picking up symptom patterns that are predictive (or anti-predictive) of rapid symptom remission, has not been performed to any significant degree.

## Conclusion

We demonstrate that generalizable predictions of treatment response can be achieved by supervised learning models trained on a small set of schizophrenia trial data. With the ongoing increases in data availability, future studies are bound to be more informative and may ultimately lead to prediction models with high clinical utility. Efforts directed at analyzing these data are, however, likely to be limited by processing power and/or software restrictions unless data sharing organizations allow for either a) offline data use or b) the purchase of more computer resources. Even then, such studies will not be as informative as they could have been had more data been available for pooling.

## Supporting information

MDAR Checklist

Supplementary figure 5 (PDF format)

Figure 1 (PDF format)

Figure 2 (PDF format)

Figure 3 (PDF format)

Supplementary figure 1 (PDF format)

Supplementary figure 2 (PDF format)

Supplementary figure 3 (PDF format)

Supplementary figure 4 (PDF format)

## Data Availability

All data can be requested from the Yale University Open Data Access Project. Accession numbers for the included studies are provided in the Supplement. The code necessary to replicate all analyses is archived with Zenodo (DOI: 10.5281/zenodo.10976022).

https://yoda.yale.edu/

https://doi.org/10.5281/zenodo.10976022

## Author contributions

Conceptualization: F.H., M.H., S.D.Ø. Methodology: F.H., M.H., A.S., S.N., S.D.Ø. Data Acquisition: F.H., S.D.Ø. Data Analysis: F.H., M.H., Visualization: F.H. Supervision: F.H., S.D.Ø. Interpretation: F.H., M.H., A.S., A.L., J.N., S.N., S.D.Ø. Writing – original draft: F.H., M.H., S.D.Ø. Writing – substantial review and editing: F.H., M.H., A.S., A.L., J.N., S.N., S.D.Ø.

## Data and materials availability

Data accession numbers for the 21 trials analyzed in this publication are provided in the SM. The code to reproduce all results in this manuscript and the supplement is available at Zenodo (DOI: 10.5281/zenodo.10976022). This study, carried out under YODA Project 2019-3941, used data obtained from the Yale University Open Data Access Project, which has an agreement with JANSSEN RESEARCH & DEVELOPMENT, L.L.C.. The interpretation and reporting of research using this data are solely the responsibility of the authors and does not necessarily represent the official views of the Yale University Open Data Access Project or JANSSEN RESEARCH & DEVELOPMENT, L.L.C.

