## Supplemental information for "The path toward generalizable clinical prediction models"

Fredrik Hieronymus *et al.*

Corresponding author: Fredrik Hieronymus


**The PDF file includes:**

Materials and methods
Tables S1 to S9
Figs. S1 to S5
Additional references

**Other Supplementary Material for this manuscript includes the following:**

MDAR Reproducibility Checklist

**Materials and methods**

*Clinical trial datasets*

Test-and-train set:

#1 – Protocol No.: PALM-JPN-4: A Randomized, Double-Blind, Placebo-Controlled, Parallel Group, Fixed-Dose, Multicenter Study of JNS010 (Paliperidone Palmitate) in Patients With Schizophrenia. NCT01299389.

#2 – Protocol No.: R076477-PSZ-3001: A Randomized, Multicenter, Double-Blind, Weight-Based, Fixed-Dose, Parallel-Group, Placebo-Controlled Study of the Efficacy and Safety of Extended Release Paliperidone for the Treatment of Schizophrenia in Adolescent Subjects, 12 to 17 Years of Age. NCT00518323. [*"Teens"* in Chekroud et al.]

#3 – Protocol No.: R076477-SCA-3001: A Randomized, Double-Blind, Placebo-Controlled, Parallel-Group, Study to Evaluate the Efficacy and Safety of Two Dosages of Paliperidone ER in the Treatment of Subjects with Schizoaffective Disorder. NCT00397033.

#4 – Protocol No.: R076477-SCA-3002: A Randomized, Double-Blind, Placebo-Controlled, Parallel-Group Study to Evaluate the Efficacy and Safety of Flexible Dose Paliperidone ER in the Treatment of Subjects with Schizoaffective Disorder. NCT00412373.

#5 – Protocol No.: R076477-SCH-3015: A Randomized, Double-Blind, Placebo-Controlled, Parallel-Group Study to Evaluate the Efficacy and Safety of Paliperidone ER Compared to Quetiapine in Subjects with an Acute Exacerbation of Schizophrenia. NCT00334126. [*"Adults, first episode"* in Chekroud et al.]

#6 – Protocol No.: R076477-SCH-302: A Randomized, 6-Week Double-Blind, Placebo-Controlled Study With an Optional 24-Week Open-Label Extension to Evaluate the Safety and Tolerability of Flexible Doses of Extended Release OROS® Paliperidone in the Treatment of Geriatric Subjects With Schizophrenia. NCT00085748. [*"Older adults"* in Chekroud et al.]

#7 – Protocol No.: R076477-SCH-303: A Randomized, Double-Blind, Placebo- and Active-Controlled, Parallel-Group, Dose-Response Study to Evaluate the Efficacy and Safety of 3 Fixed Dosages of Extended Release OROS® Paliperidone (6, 9, and 12 mg/day) and Olanzapine (10 mg/day), With Open-Label Extension, in the Treatment of Subjects With Schizophrenia. NCT00078039. [*"Adults – Chronic #2"* in Chekroud et al.]

#8 – Protocol No.: R076477-SCH-304: A Randomized, Double-Blind, Placebo- and Active-Controlled, Parallel-Group, Dose-Response Study to Evaluate the Efficacy and Safety of 2 Fixed Dosages of Extended Release OROS® Paliperidone (6 and 12 mg/day) and Olanzapine (10 mg/day), With Open-Label Extension, in the Treatment of Subjects With Schizophrenia. NCT00077714.

#9 – Protocol No.: R076477-SCH-305: A Randomized, Double-Blind, Placebo- and Active-Controlled, Parallel-Group, Dose-Response Study to Evaluate the Efficacy and Safety of 3 Fixed Dosages of Extended Release OROS® Paliperidone (3, 9, and 15 mg/day) and Olanzapine (10 mg/day), With Open-Label Extension, in the Treatment of Subjects With Schizophrenia. NCT00083668. [*"Adults – Chronic #1"* in Chekroud et al.]

#10 – Protocol No.: R076477-SCH-4012: A Randomized, Double-Blind, Placebo- and Active-Controlled, Parallel-Group Study to Evaluate the Efficacy and Safety of a Fixed Dosage of 1.5 mg/day of Paliperidone Extended Release (ER) in the Treatment of Subjects With Schizophrenia. NCT00524043.

#11 – Protocol No.: R092670-PSY-3003: A Randomized, Double-Blind, Placebo-Controlled, Parallel-Group, Dose-Response Study to Evaluate the Efficacy and Safety of 3 Fixed Doses (50 mg eq., 100 mg eq., and 150 mg eq.) of Paliperidone Palmitate in Subjects With Schizophrenia. NCT00210548.

#12 – Protocol No.: R092670-PSY-3004: A Randomized, Double-Blind, Placebo-Controlled, Parallel-Group, Dose-Response Study to Evaluate the Efficacy and Safety of 3 Fixed Doses (25 mg eq., 50 mg eq., and 100 mg eq.) of Paliperidone Palmitate in Subjects With Schizophrenia. NCT00101634.

#13 – Protocol No.: R092670-PSY-3007: A Randomized, Double-Blind, Placebo-Controlled, Parallel-Group, Dose-Response Study to Evaluate the Efficacy and Safety of 3 Fixed Doses (25 mg eq., 100 mg eq., and 150 mg eq.) of Paliperidone Palmitate in Subjects With Schizophrenia. NCT00590577.

#14 – Protocol No.: R092670-SCH-201: A Randomized, Double-Blind, Placebo-Controlled Study to Evaluate the Efficacy and Safety of 50 and 100 mg eq. of Paliperidone Palmitate in Subjects With Schizophrenia. NCT00074477.

#15 – Protocol No.: RIS-INT-3: A randomized, double-blind, placebo-controlled multicenter study compared four fixed doses of risperidone and one dose of haloperidol in schizophrenic patients. Patients were treated for eight weeks with risperidone 2, 6, 10, or 16 mg, haloperidol 20 mg, or placebo. NCT00249132.

#16 – Protocol No.: RIS-SCH-302: A Randomized, Double-Blind, Placebo-Controlled Clinical Study of the Efficacy and Safety of Risperidone for the Treatment of Schizophrenia in Adolescents. NCT00088075.

#17 – Protocol No.: RIS-USA-121: Risperidone depot (microspheres) vs. placebo in the treatment of patients with schizophrenia. NCT00253136.

#18 – Protocol No.: RIS-USA-72: The Safety and Effectiveness of Risperidone 8 mg QD and 4 mg QD Compared to Placebo in the Treatment of Schizophrenia. No NCT number available.

<u>Extender set:</u>

Protocol No.: R092670-PSY-3002: A Randomized, Double Blind, Parallel-Group Comparative Study of Flexibly Dosed Paliperidone Palmitate (25, 50, 75, or 100 mg eq.) Administered Every 4 Weeks and Flexibly Dosed RISPERDAL® CONSTA® (25, 37.5, or 50 mg) Administered Every 2 Weeks in Subjects With Schizophrenia. NCT00210717.

Protocol No.: R092670-PSY-3006: A Randomized, Double-Blind, Parallel-Group, Comparative Study of Flexible Doses of Paliperidone Palmitate and Flexible Doses of Risperidone Long-Acting Intramuscular Injection in Subjects With Schizophrenia. NCT00589914

Protocol No.: RIS-USA-231: The Efficacy and Safety of Risperidone in the Treatment of Adolescents with Schizophrenia. NCT00034749.

Data accuracy and completeness was verified by comparing the provided data to that reported in study reports provided by YODA and with those available in public reports from the United States Food and Drug Administration,[1-7] the European Medicines Agency,[8] and ClinicalTrials.gov[9-11]

*Outcomes*

Symptomatic remission as defined by the Remission in Schizophrenia Working Group was used as the outcome for all analyses. For symptomatic remission to be at hand the patient must have a Positive and Negative Syndrome Scale (PANSS)[12] score of no more than three on three positive symptoms (P01 Delusions, P02 Conceptual disorganization and P03 Hallucinations), three negative symptoms (N01 Blunted affect, N04 Passive/apathetic social withdrawal, and N06 Lack of spontaneity and flow of conversation) and two general symptoms (G05 Mannerisms and posturing and G09 Unusual thought content).

*Analyses*

For simplicity we use the same subheadings here as in the main manuscript.

<u>Contrasting single study and random subsample response prediction estimates</u>

We replicated the analyses by Chekroud using random subsampling from a larger schizophrenia data set to assess a) whether their observed accuracy was compatible with random draws of the same number of patients and b) whether their results were negatively impacted by the inclusion of a high number of predictors (p=217 for elastic net models and 137 for random forest models).

The data set used by Chekroud and colleagues consisted of four week completer data (with some data imputation) for 1513 patients spread across five placebo-controlled trials (n=99, 182, 321, 430, 481). Our analysis set contained 4634 patients with completer data spread across eighteen placebo-controlled trials. Since there was very little missing data for the predictors we included (~0.5% for all predictors combined), we did not do any data imputation but instead opted to drop the 25 cases for which we lacked data on at least one predictor.

To assess the impact of including a high number of predictors, we focused our analyses on a reduced predictor set of 33 variables: age, sex, treatment (antipsychotic or placebo) and baseline scores on the 30 items of the Positive and Negative Syndrome Scale (PANSS-30). These predictors were included also in the analyses by Chekroud et al and we refer to them as the *sparse* predictor set. We then built a second *noisy* predictor set which included the *sparse* predictor set as well as several simulated predictors known to be uninformative (i.e., uncorrelated to the studied outcome). This was done to assess the models' ability to parse out uninformative data. To facilitate comparison, we included as many uninformative predictors as the difference between our *sparse* predictor set and the predictor sets used by Chekroud and colleagues (i.e., p=184 and 104, for elastic net and random forest models, respectively). To resemble the different types of data that can be encountered in a clinical data set, the noise variables were simulated with one quarter of the variables each having an exponential, normal, Poisson (lambda = 5) and uniform distribution, respectively. Noise variables were simulated using the corresponding built-in R functions (rexp, rnorm, rpois and runif).

To estimate the variance inherent in the data we used a Monte Carlo subsampling method on the *test-and-train* set to construct training sets of the same five sizes as used by Chekroud and colleagues to estimate out-of-sample balanced accuracy (i.e., 1513 minus the number of patients in each holdout trial). Subsampling was done 50 times for each trial for the elastic net model and, due to processing power limitations, 25 times for each trial for the random forest models. We, like Chekroud, trained each elastic net model using 400 combinations of alpha and lambda values according to carets built in-tuning algorithm, and each random forest model on 20 different mtry values also using carets prespecified tunings. Caret's preprocessing function was used to center and scale all variables as well as to remove zero- and near zero-variance variables.

For each such run we then used Monte Carlo subsampling to randomly draw a test set of equal size to the corresponding holdout trial (i.e., so that each training and test set size added up to 1513 patients) from the cases not selected for training. We then used the fitted models to predict symptomatic remission. This process was repeated (using non-parametric bootstrap) 100 times for each Monte Carlo subsampling. In total, each out-of-sample balanced accuracy estimate is thus based on 5000 sub- and resamples for the elastic net models, and 2500 sub- and resamples for the random forest model.

<u>Feature selection performance of elastic net in simulated data with weak predictors</u>

We simulated data sets ranging in size from 2000 to 25000 cases (2000, 3000, 4000, 5000, 7500, 10000, 15000, 25000). Cases were simulated by a vector Y of random normal variables ($\mu = 0$, $\sigma = 1$) with cases defined as Y > 0. Each data set included 50 simulated weak predictors with correlations (r) of 0.05, 0.10 or 0.15, respectively, to the case variable. Weak predictors were generated using the formula:

$$X_i = rY + \sqrt{1 - r^2} \cdot Z_i$$

where Y is the case variable vector and $Z_i$ are independent random normal vectors with $\mu = 0$ and $\sigma = 1$. Each data set also included a variable number (p=150 or 450) of random normal noise variables ($\mu = 0$, $\sigma = 1$); corresponding to 3:1 and 9:1, respectively, ratios of uninformative to informative variables. Each simulation was repeated 50 times to get more stable estimates.

In total, we thus simulated 2400 (8 · 50 · 3 · 2) data sets of different sizes and with differing information content and signal-to-noise ratios. The performance of a) an elastic net model tuned using the same built-in 400 hyperparameter combinations as above and b) a generalized linear model, was evaluated for all data sets. Model performance was compared to the balanced accuracy (BAC) given by the sum (W) of all n informative predictors (i.e., disregarding all uninformative variables):

$$W = X_1 + X_2 + \cdots X_n$$

If we use W > 0 as cutoff to maximize BAC and then condition on case status Y and define $W_+ = W|Y > 0$ and $W_- = W|Y < 0$, then the expectation, variance and sensitivity are

$$\mu W_+ = E(W_+) = nr\sqrt{2/\pi}$$

$$\sigma_{W_+}^2 = Var(W_+) = n^2 r^2 \left(1 - \frac{2}{\pi}\right) + n(1 - r^2)$$

$$sensitivity = P(W > 0|Y > 0) \approx \Phi(\frac{\mu W_+}{\sigma_{W_+}})$$

where $\Phi$ is the cumulative distribution function of a standardized normal distribution. By symmetry, $\mu W_- = -\mu W_+,\ \sigma^2_{W_+} = \sigma^2_{W_-}$ and $specificity = sensitivity$, thus:

$$BAC = \frac{sensitivity + specificity}{2} \approx \Phi(\frac{\mu W_+}{\sigma_{W_+}})$$

Ensemble prediction

Five classification models: bagged classification and regression training (Bagged CART; caret package: treebag), elastic net (caret package: glmnet), generalized linear model (caret package: glm), random forest (caret package: rf), and extreme gradient boosting trees (caret package: xgbTree) were included. The glm and treebag models do not require hyperparameter tuning, the rf model has a finite hyperparameter grid where all possible parameters can be explored using grid search, and glmnet and xgbTree can only be partially explored using semi-random search strategies.

Using the full *test-and-train* set, we applied a grid search to find the best hyperparameters for the rf algorithm, and we sought for the optimal hyperparameter tuning by using genetic algorithms (R-package: GA). The search bounds were iteratively tightened, first after an initial run of ten generations, and then after sequential runs of 100 generations in cases where there was still improvement occurring at least every 30th generation. The best solution from each run was included as a suggestion in subsequent runs. For all runs, the mutation rate was set to 0.25 and the crossover probability to 0.80, with a population size of 100. All runs except the first one were set to stop if there was no improvement after 30 generations. Local optimization was used initially but since it was very resource intensive and did not seem to result in any additional improvement it was only used for the initial runs. Additional parameters as well as the outcomes for all searches are detailed in Supplementary table 1. We also tried Bayesian Optimization (R-package: rBayesianOptimization), but this did not yield better results (data not shown).

We then used the obtained hyperparameters and constructed ensembles of models in caret using linear greedy optimization. The ensembles were trained on Monte Carlo subsampled test-train splits. Test set size was fixed at 250 cases while training sets varied in size between 384 and 4384 cases (384, 884, 1384, 2384, 3384 and 4384). The process was repeated 50 times for each training set size.  For the largest training set size – which utilized all cases – the training set was bootstrapped, rather than subsampled, 50 times. The test sets were similarly bootstrapped 100 times yielding in total 5000 estimates for each training set size. Performance was also tested against the *extender* set for each subsampled training set.

To test between-study generalizability, we also did out-of-sample predictions by training the ensemble models on all data except one study and using that study as the test set. To obtain stable estimates, the test sets were bootstrapped 100 times. To allow for comparisons against random chance, we also did 50 Monte Carlo subsamples of training and test sets corresponding in size to the smallest and largest study, respectively. Test sets were again bootstrapped 100 times. These two data points were used to create 95% CLs for expected balance accuracy.

**Supplementary table 1 – Hyperparameter tuning using genetic algorithms and grid search**

| Model | Run | Max iterations | Elitism | Bounds | Best tuning | Outcome |
|---|---|---|---|---|---|---|
| glmnet | 1 | 10 | 2 | alpha: 0 to 1<br>lambda: 0 to 0.2 | alpha: 0.0218092<br>lambda: 0.0569022 | AUC-ROC = 70.66% |
| glmnet | 2 | 100 | 5 | alpha: 0 to 0.060<br>lambda: 0 to 0.100 | alpha: 0.0205956<br>lambda: 0.0669554 | AUC-ROC = 70.74%<br>Stopped after 84 iterations |
| xgbTree | 1 | 10 | 2 | eta: 0.01 to 0.15<br>depth: 2 to 8<br>colsample_bytree: 0.2 to 0.8<br>subsample: 0.2 to 0.8<br>nrounds: 100 to 700<br>gamma = 0<br>min_child_weight = 5 | eta: 0.0213504<br>depth: 5<br>colsample_bytree: 0.343392<br>subsample: 0.560772<br>nrounds: 273<br>gamma = 0<br>min_child_weight = 5 | AUC-ROC = 73.66% |
| xgbTree | 2 | 100 | 5 | eta: 0.01 to 0.04<br>depth: 2 to 5<br>colsample_bytree: 0.3 to 0.7<br>subsample: 0.3 to 0.7<br>nrounds: 250 to 400<br>gamma = 0<br>min_child_weight = 5 | eta: 0.0207611<br>depth: 5<br>colsample_bytree: 0.347036<br>subsample: 0.504784<br>nrounds: 348<br>gamma = 0<br>min_child_weight = 5 | AUC-ROC = 73.85%<br>Stopped after 44 iterations |
| RF | N/A | N/A | N/A | mtry=2 to 33 | mtry=3 | AUC-ROC = 73.62%<br>Grid search |

*The final values reported under "Best tuning" for each model are the hyperparameters used for all ensemble predictions. The glm and treebag models do not use tunable hyperparameters.*

**Supplementary table 2 – Baseline demographics for all studies**

| Dataset | Protocol No. | N completers | N placebo (%) | N male (%) | Age (mean) | Age (sd) |
|---|---|---|---|---|---|---|
| **Test-and-train** | PALM-JPN-4 (#1) | 222 | 106 (48) | 127 (57) | 45.3 | 13.2 |
| | R076477-PSZ-3001 (#2) | 156 | 32 (21) | 93 (60) | 15.2 | 1.5 |
| | R076477-SCA-3001 (#3) | 215 | 63 (29) | 142 (66) | 37.0 | 9.9 |
| | R076477-SCA-3002 (#4) | 197 | 53 (27) | 106 (54) | 37.6 | 9.0 |
| | R076477-SCH-3015 (#5) | 323 | 60 (19) | 207 (64) | 35.8 | 10.8 |
| | R076477-SCH-302 (#6) | 98 | 30 (31) | 27 (28) | 69.5 | 4.5 |
| | R076477-SCH-303 (#7) | 471 | 74 (16) | 241 (51) | 37.0 | 11.3 |
| | R076477-SCH-304 (#8) | 231 | 48 (21) | 169 (73) | 42.3 | 10.7 |
| | R076477-SCH-305 (#9) | 427 | 66 (15) | 286 (67) | 37.3 | 10.7 |
| | R076477-SCH-4012 (#10) | 124 | 39 (31) | 85 (69) | 40.8 | 12.3 |
| | R092670-PSY-3003 (#11) | 230 | 81 (35) | 156 (68) | 39.2 | 10.6 |
| | R092670-PSY-3004 (#12) | 371 | 84 (23) | 245 (66) | 39.8 | 11.4 |
| | R092670-PSY-3007 (#13) | 469 | 107 (23) | 303 (65) | 39.1 | 10.8 |
| | R092670-SCH-201 (#14) | 172 | 49 (28) | 111 (65) | 39.1 | 10.5 |
| | RIS-INT-3 (#15) | 336 | 44 (13) | 284 (85) | 37.6 | 10.4 |
| | SCH-302 (#16) | 137 | 47 (34) | 88 (64) | 15.7 | 1.3 |
| | USA-121 (#17) | 294 | 62 (21) | 205 (70) | 39.1 | 9.6 |
| | USA-72 (#18) | 161 | 52 (32) | 129 (80) | 37.7 | 9.1 |
| **Extender** | USA-231 | 198 | 0 (0) | 110 (56) | 15.2 | 1.9 |
| | R092670-PSY-3006 | 1070 | 0 (0) | 610 (57) | 38.9 | 12.0 |
| | R076477-PSZ-3003 | 195 | 0 (0) | 130 (67) | 15.3 | 1.5 |

**Supplementary table 3 – Baseline item scores for the *test-and-train* set and the *extender* set.**

| | Test-and-train set | | Extender set | |
|---|---|---|---|---|
| **Item** | **Mean** | **SD** | **Mean** | **SD** |
| P1: Delusions | 4.06 | 1.17 | 3.61 | 1.09 |
| P2: Conceptual disorganization | 3.53 | 1.16 | 3.38 | 0.99 |
| P3: Hallucinations | 3.61 | 1.43 | 3.12 | 1.39 |
| P4: Excitement | 2.85 | 1.21 | 2.54 | 1.11 |
| P5: Grandiosity | 2.38 | 1.39 | 2.00 | 1.11 |
| P6: Suspiciousness/persecution | 3.84 | 1.18 | 3.56 | 1.09 |
| P7: Hostility | 2.46 | 1.24 | 2.13 | 1.05 |
| N1: Blunted affect | 3.40 | 1.18 | 3.47 | 1.04 |
| N2: Emotional withdrawal | 3.54 | 1.06 | 3.50 | 0.93 |
| N3: Poor rapport | 2.97 | 1.13 | 3.01 | 1.03 |
| N4: Passive/apathetic social withdrawal | 3.56 | 1.14 | 3.56 | 1.05 |
| N5: Difficulty in abstract thinking | 3.80 | 1.20 | 3.48 | 1.07 |
| N6: Lack of spontaneity and flow of conversation | 3.06 | 1.26 | 3.11 | 1.12 |
| N7: Stereotyped thinking | 3.12 | 1.11 | 3.04 | 1.01 |
| G1: Somatic concern | 2.52 | 1.24 | 2.29 | 1.14 |
| G2: Anxiety | 3.16 | 1.11 | 2.99 | 1.02 |
| G3: Guilt feelings | 2.14 | 1.23 | 1.88 | 1.01 |
| G4: Tension | 3.07 | 1.09 | 2.88 | 0.92 |
| G5: Mannerisms and posturing | 2.47 | 1.16 | 2.38 | 1.13 |
| G6: Depression | 2.57 | 1.28 | 2.33 | 1.11 |
| G7: Motor retardation | 2.35 | 1.15 | 2.37 | 1.09 |
| G8: Uncooperativeness | 2.29 | 1.20 | 2.10 | 1.10 |
| G9: Unusual thought content | 3.50 | 1.18 | 3.12 | 1.12 |
| G10: Disorientation | 2.02 | 1.07 | 1.88 | 0.95 |
| G11: Poor attention | 2.94 | 1.09 | 3.00 | 0.93 |
| G12: Lack of judgment and insight | 3.69 | 1.16 | 3.47 | 1.03 |
| G13: Disturbance of volition | 3.03 | 1.08 | 3.09 | 1.02 |
| G14: Poor impulse control | 2.53 | 1.18 | 2.26 | 1.09 |
| G15: Preoccupation | 3.32 | 1.11 | 3.14 | 0.97 |
| G16: Active social avoidance | 3.35 | 1.10 | 3.26 | 1.03 |

**Supplementary table 4 – Additional metrics for the ensemble model**

| Sample | Training set size | Train BAC (mean) | Train BAC (sd) | Test BAC (mean) | Test BAC (sd) | False positives (mean) | True positives (mean) | False negatives (mean) | True negatives (mean) |
|---|---|---|---|---|---|---|---|---|---|
| **Subsamples** | 384 | 0.599 | 0.034 | 0.599 | 0.033 | 22 | 29 | 59 | 141 |
| | 884 | 0.616 | 0.019 | 0.618 | 0.030 | 21 | 32 | 56 | 141 |
| | 1384 | 0.627 | 0.014 | 0.628 | 0.029 | 21 | 34 | 54 | 141 |
| | 2384 | 0.634 | 0.010 | 0.635 | 0.030 | 22 | 35 | 52 | 141 |
| | 3384 | 0.637 | 0.007 | 0.638 | 0.028 | 21 | 36 | 52 | 141 |
| | 4384 | 0.640 | 0.003 | 0.640 | 0.039 | 22 | 37 | 52 | 139 |
| **Extender** | 384 | N/A | N/A | 0.628 | 0.030 | 146 | 268 | 355 | 694 |
| | 884 | N/A | N/A | 0.656 | 0.018 | 164 | 316 | 307 | 676 |
| | 1384 | N/A | N/A | 0.661 | 0.018 | 164 | 323 | 301 | 675 |
| | 2384 | N/A | N/A | 0.668 | 0.014 | 170 | 335 | 288 | 671 |
| | 3384 | N/A | N/A | 0.667 | 0.014 | 171 | 335 | 288 | 669 |
| | 4384 | N/A | N/A | 0.668 | 0.013 | 169 | 336 | 288 | 670 |

*The figures are averaged over 50 runs for each train set size and then rounded. The number of true and false positives and negatives may hence not always line up with the size of the test (n=250) and extender (n=1413) sets.*

**Supplementary table 5 – Additional metrics for the glm model**

| Sample | Training set size | Train BAC (mean) | Train BAC (sd) | Test BAC (mean) | Test BAC (sd) | False positives (mean) | True positives (mean) | False negatives (mean) | True negatives (mean) |
|---|---|---|---|---|---|---|---|---|---|
| **Subsamples** | 384 | 0.586 | 0.029 | 0.589 | 0.032 | 31 | 33 | 55 | 131 |
| | 884 | 0.600 | 0.023 | 0.601 | 0.029 | 25 | 31 | 57 | 137 |
| | 1384 | 0.605 | 0.014 | 0.605 | 0.029 | 23 | 31 | 57 | 139 |
| | 2384 | 0.607 | 0.012 | 0.608 | 0.029 | 22 | 31 | 57 | 140 |
| | 3384 | 0.609 | 0.006 | 0.607 | 0.028 | 21 | 30 | 58 | 141 |
| | 4384 | 0.610 | 0.003 | 0.605 | 0.040 | 21 | 30 | 59 | 140 |
| **Extender** | 384 | N/A | N/A | 0.608 | 0.026 | 229 | 304 | 319 | 611 |
| | 884 | N/A | N/A | 0.631 | 0.023 | 205 | 315 | 307 | 635 |
| | 1384 | N/A | N/A | 0.633 | 0.020 | 187 | 304 | 319 | 653 |
| | 2384 | N/A | N/A | 0.643 | 0.016 | 182 | 313 | 310 | 658 |
| | 3384 | N/A | N/A | 0.644 | 0.014 | 179 | 312 | 311 | 661 |
| | 4384 | N/A | N/A | 0.644 | 0.013 | 174 | 309 | 315 | 665 |

*The figures are averaged over 50 runs for each train set size and then rounded. The number of true and false positives and negatives may hence not always line up with the size of the test (n=250) and extender (n=1413) sets.*

**Supplementary table 6 – Additional metrics for the glmnet model**

| Sample | Training set size | Train BAC (mean) | Train BAC (sd) | Test BAC (mean) | Test BAC (sd) | False positives (mean) | True positives (mean) | False negatives (mean) | True negatives (mean) |
|---|---|---|---|---|---|---|---|---|---|
| **Subsamples** | 384 | 0.583 | 0.030 | 0.584 | 0.031 | 22 | 26 | 61 | 140 |
| | 884 | 0.588 | 0.021 | 0.589 | 0.028 | 18 | 25 | 63 | 144 |
| | 1384 | 0.592 | 0.016 | 0.591 | 0.027 | 17 | 25 | 63 | 145 |
| | 2384 | 0.593 | 0.012 | 0.593 | 0.027 | 16 | 25 | 62 | 146 |
| | 3384 | 0.594 | 0.006 | 0.592 | 0.026 | 16 | 25 | 63 | 146 |
| | 4384 | 0.593 | 0.002 | 0.590 | 0.036 | 16 | 25 | 65 | 145 |
| **Extender** | 384 | N/A | N/A | 0.606 | 0.026 | 155 | 247 | 376 | 685 |
| | 884 | N/A | N/A | 0.622 | 0.024 | 138 | 255 | 368 | 702 |
| | 1384 | N/A | N/A | 0.619 | 0.020 | 126 | 242 | 381 | 714 |
| | 2384 | N/A | N/A | 0.626 | 0.016 | 126 | 251 | 372 | 714 |
| | 3384 | N/A | N/A | 0.627 | 0.013 | 122 | 248 | 375 | 718 |
| | 4384 | N/A | N/A | 0.623 | 0.012 | 121 | 243 | 380 | 719 |

*The figures are averaged over 50 runs for each train set size and then rounded. The number of true and false positives and negatives may hence not always line up with the size of the test (n=250) and extender (n=1413) sets.*

**Supplementary table 7 – Additional metrics for the rf model**

| Sample | Training set size | Train BAC (mean) | Train BAC (sd) | Test BAC (mean) | Test BAC (sd) | False positives (mean) | True positives (mean) | False negatives (mean) | True negatives (mean) |
|---|---|---|---|---|---|---|---|---|---|
| **Subsamples** | 384 | 0.590 | 0.032 | 0.591 | 0.033 | 17 | 25 | 63 | 146 |
| | 884 | 0.603 | 0.018 | 0.604 | 0.029 | 16 | 27 | 61 | 146 |
| | 1384 | 0.610 | 0.014 | 0.612 | 0.028 | 16 | 28 | 60 | 146 |
| | 2384 | 0.617 | 0.010 | 0.618 | 0.028 | 16 | 29 | 58 | 146 |
| | 3384 | 0.619 | 0.007 | 0.620 | 0.026 | 15 | 29 | 59 | 147 |
| | 4384 | 0.622 | 0.003 | 0.621 | 0.036 | 16 | 31 | 59 | 144 |
| **Extender** | 384 | N/A | N/A | 0.625 | 0.029 | 117 | 243 | 380 | 723 |
| | 884 | N/A | N/A | 0.645 | 0.019 | 127 | 275 | 348 | 713 |
| | 1384 | N/A | N/A | 0.647 | 0.019 | 120 | 272 | 351 | 719 |
| | 2384 | N/A | N/A | 0.655 | 0.015 | 121 | 283 | 340 | 719 |
| | 3384 | N/A | N/A | 0.654 | 0.014 | 120 | 281 | 342 | 720 |
| | 4384 | N/A | N/A | 0.655 | 0.013 | 118 | 282 | 342 | 721 |

*The figures are averaged over 50 runs for each train set size and then rounded. The number of true and false positives and negatives may hence not always line up with the size of the test (n=250) and extender (n=1413) sets.*

**Supplementary table 8 – Additional metrics for the treebag model**

| Sample | Training set size | Train BAC (mean) | Train BAC (sd) | Test BAC (mean) | Test BAC (sd) | False positives (mean) | True positives (mean) | False negatives (mean) | True negatives (mean) |
|---|---|---|---|---|---|---|---|---|---|
| **Subsamples** | 384 | 0.593 | 0.030 | 0.596 | 0.032 | 31 | 33 | 54 | 132 |
| | 884 | 0.604 | 0.017 | 0.607 | 0.031 | 28 | 34 | 54 | 134 |
| | 1384 | 0.613 | 0.013 | 0.613 | 0.030 | 27 | 35 | 53 | 135 |
| | 2384 | 0.619 | 0.012 | 0.617 | 0.030 | 27 | 35 | 53 | 136 |
| | 3384 | 0.620 | 0.009 | 0.619 | 0.029 | 26 | 35 | 53 | 136 |
| | 4384 | 0.621 | 0.005 | 0.625 | 0.040 | 25 | 37 | 53 | 135 |
| **Extender** | 384 | N/A | N/A | 0.621 | 0.027 | 196 | 296 | 327 | 644 |
| | 884 | N/A | N/A | 0.632 | 0.020 | 197 | 310 | 313 | 643 |
| | 1384 | N/A | N/A | 0.631 | 0.019 | 188 | 303 | 321 | 652 |
| | 2384 | N/A | N/A | 0.636 | 0.016 | 194 | 313 | 310 | 646 |
| | 3384 | N/A | N/A | 0.636 | 0.017 | 190 | 311 | 312 | 649 |
| | 4384 | N/A | N/A | 0.635 | 0.015 | 186 | 307 | 316 | 653 |

*The figures are averaged over 50 runs for each train set size and then rounded. The number of true and false positives and negatives may hence not always line up with the size of the test (n=250) and extender (n=1413) sets.*

**Supplementary table 9 – Additional metrics for the xgbTree model**

| Sample | Training set size | Train BAC (mean) | Train BAC (sd) | Test BAC (mean) | Test BAC (sd) | False positives (mean) | True positives (mean) | False negatives (mean) | True negatives (mean) |
|---|---|---|---|---|---|---|---|---|---|
| **Subsamples** | 384 | 0.601 | 0.035 | 0.605 | 0.033 | 25 | 32 | 56 | 137 |
| | 884 | 0.616 | 0.015 | 0.619 | 0.030 | 23 | 34 | 54 | 139 |
| | 1384 | 0.627 | 0.013 | 0.627 | 0.029 | 23 | 35 | 53 | 139 |
| | 2384 | 0.633 | 0.011 | 0.632 | 0.029 | 22 | 35 | 53 | 140 |
| | 3384 | 0.635 | 0.007 | 0.636 | 0.028 | 21 | 35 | 53 | 141 |
| | 4384 | 0.636 | 0.004 | 0.633 | 0.041 | 21 | 36 | 54 | 139 |
| **Extender** | 384 | N/A | N/A | 0.641 | 0.022 | 176 | 306 | 317 | 664 |
| | 884 | N/A | N/A | 0.653 | 0.018 | 190 | 331 | 292 | 651 |
| | 1384 | N/A | N/A | 0.657 | 0.018 | 177 | 327 | 297 | 663 |
| | 2384 | N/A | N/A | 0.662 | 0.015 | 177 | 333 | 290 | 664 |
| | 3384 | N/A | N/A | 0.664 | 0.015 | 173 | 332 | 291 | 667 |
| | 4384 | N/A | N/A | 0.665 | 0.013 | 168 | 331 | 293 | 672 |

*The figures are averaged over 50 runs for each train set size and then rounded. The number of true and false positives and negatives may hence not always line up with the size of the test (n=250) and extender (n=1413) sets.*

**Supplementary figure 1 – Four-week remission: BAC for glm (subsampled test data, extender test data, and single-study data)**

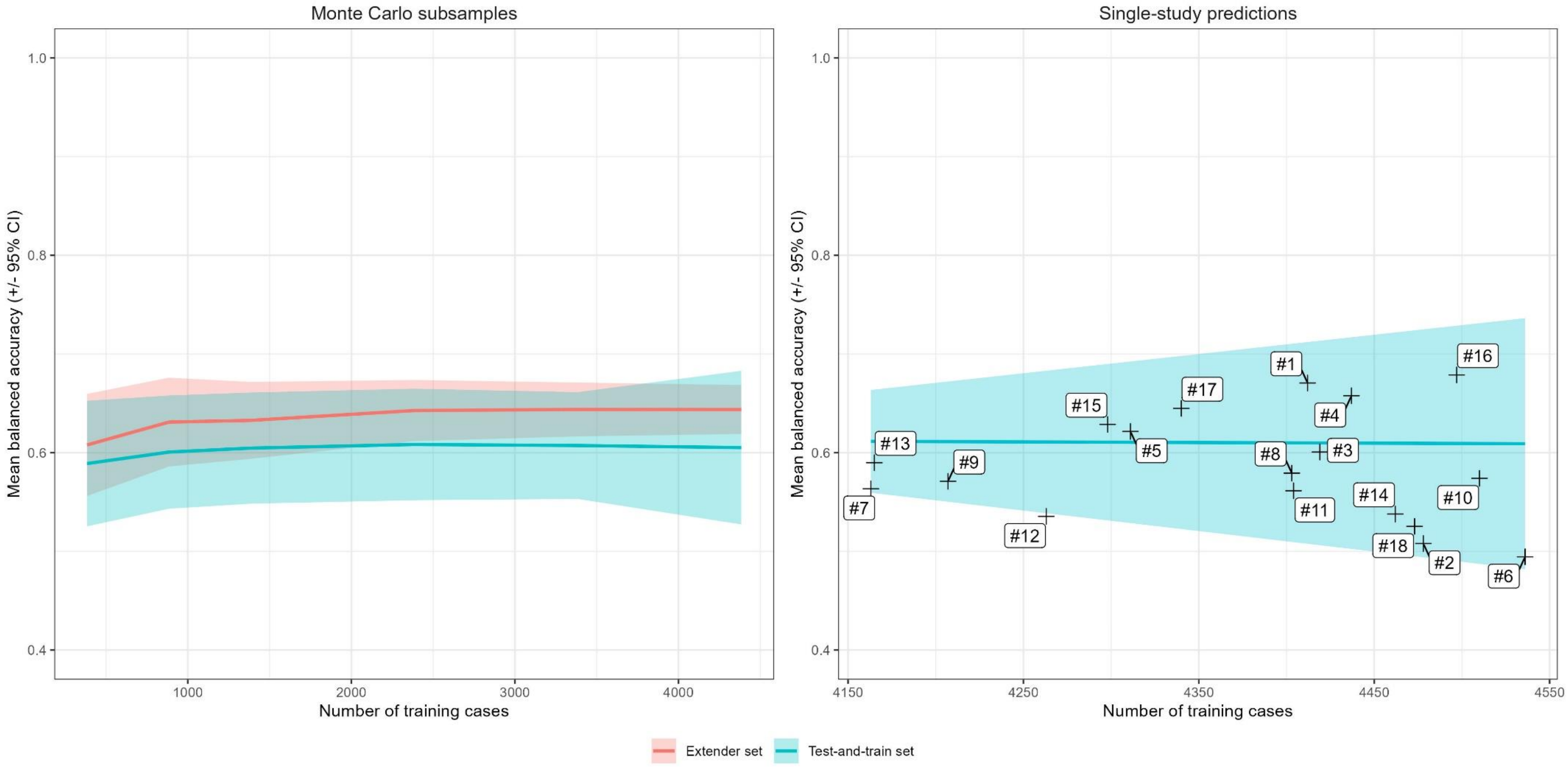


*The left panel gives mean and 95% confidence interval (CI) BAC for subsampled training sets ranging in size from 384 to 4384 cases. The right panel shows the same for subsampled and single-study BAC for leave-one-study-out analyses. Each number #1 to #18 represents one study in the train-and-test set (see Test-and-train set under **Materials and methods**).*

**Supplementary figure 2 – Four-week remission: BAC for glmnet (subsampled test data, extender test data, and single-study data)**

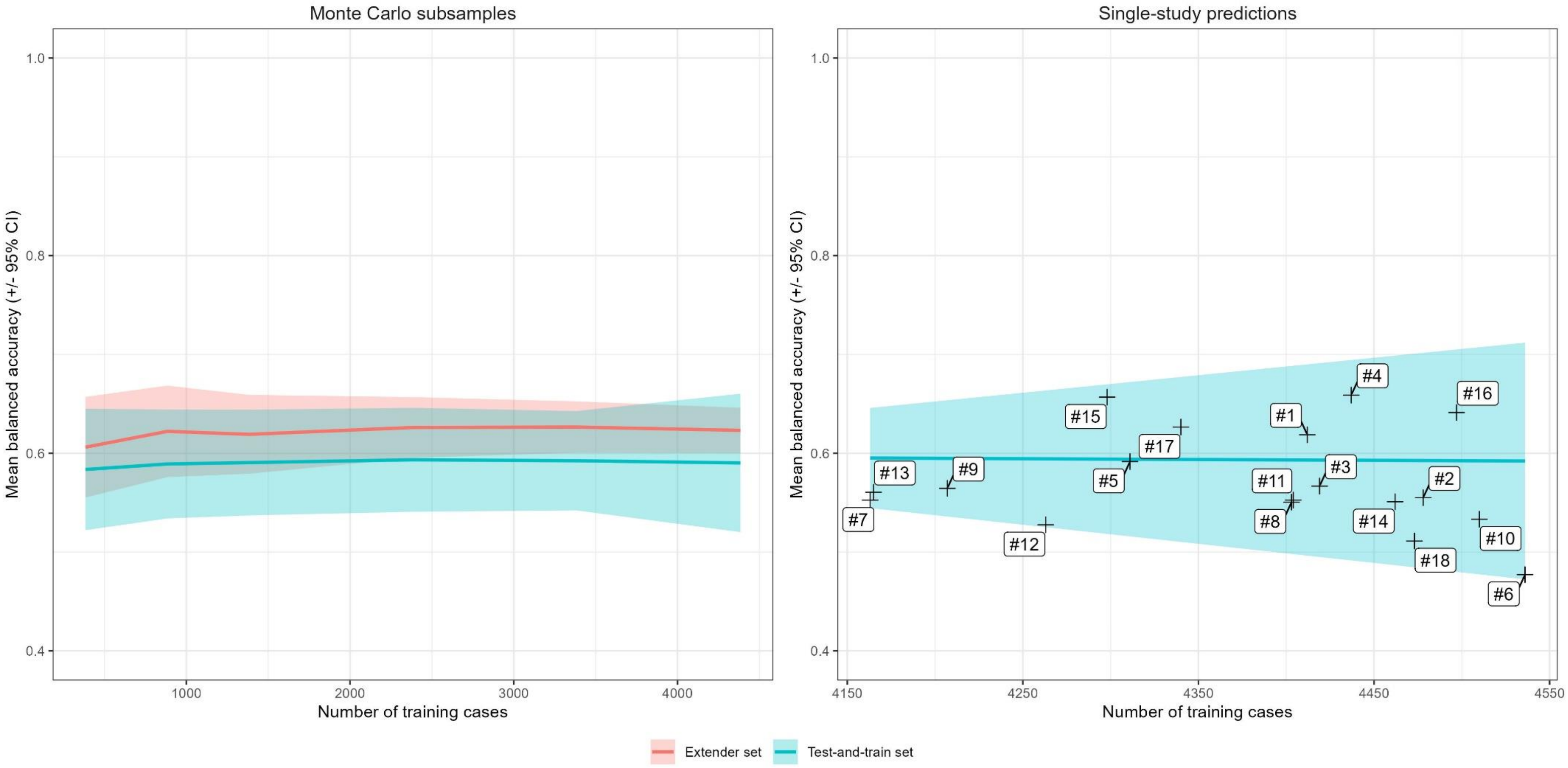


*The left panel gives mean and 95% confidence interval (CI) BAC for subsampled training sets ranging in size from 384 to 4384 cases. The right panel shows the same for subsampled and single-study BAC for leave-one-study-out analyses. Each number #1 to #18 represents one study in the train-and-test set (see Test-and-train set under* ***Materials and methods****).*

**Supplementary figure 3 – Four-week remission: BAC for rf (subsampled test data, extender test data, and single-study data)**

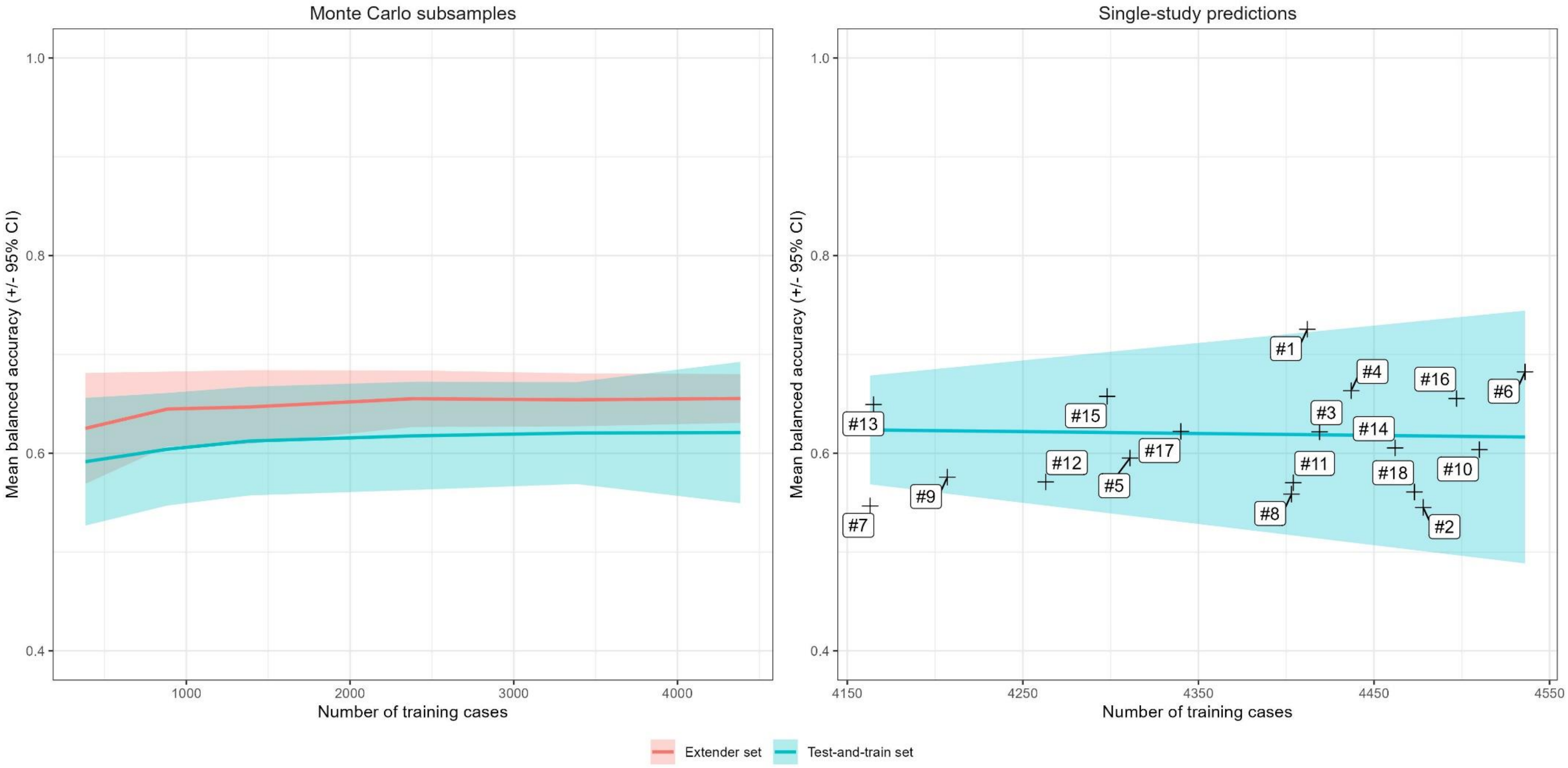


*The left panel gives mean and 95% confidence interval (CI) BAC for subsampled training sets ranging in size from 384 to 4384 cases. The right panel shows the same for subsampled and single-study BAC for leave-one-study-out analyses. Each number #1 to #18 represents one study in the train-and-test set (see Test-and-train set under* ***Materials and methods****).*

**Supplementary figure 4 – BAC for treebag model (subsampled test data, extender test data, and single-study data)**

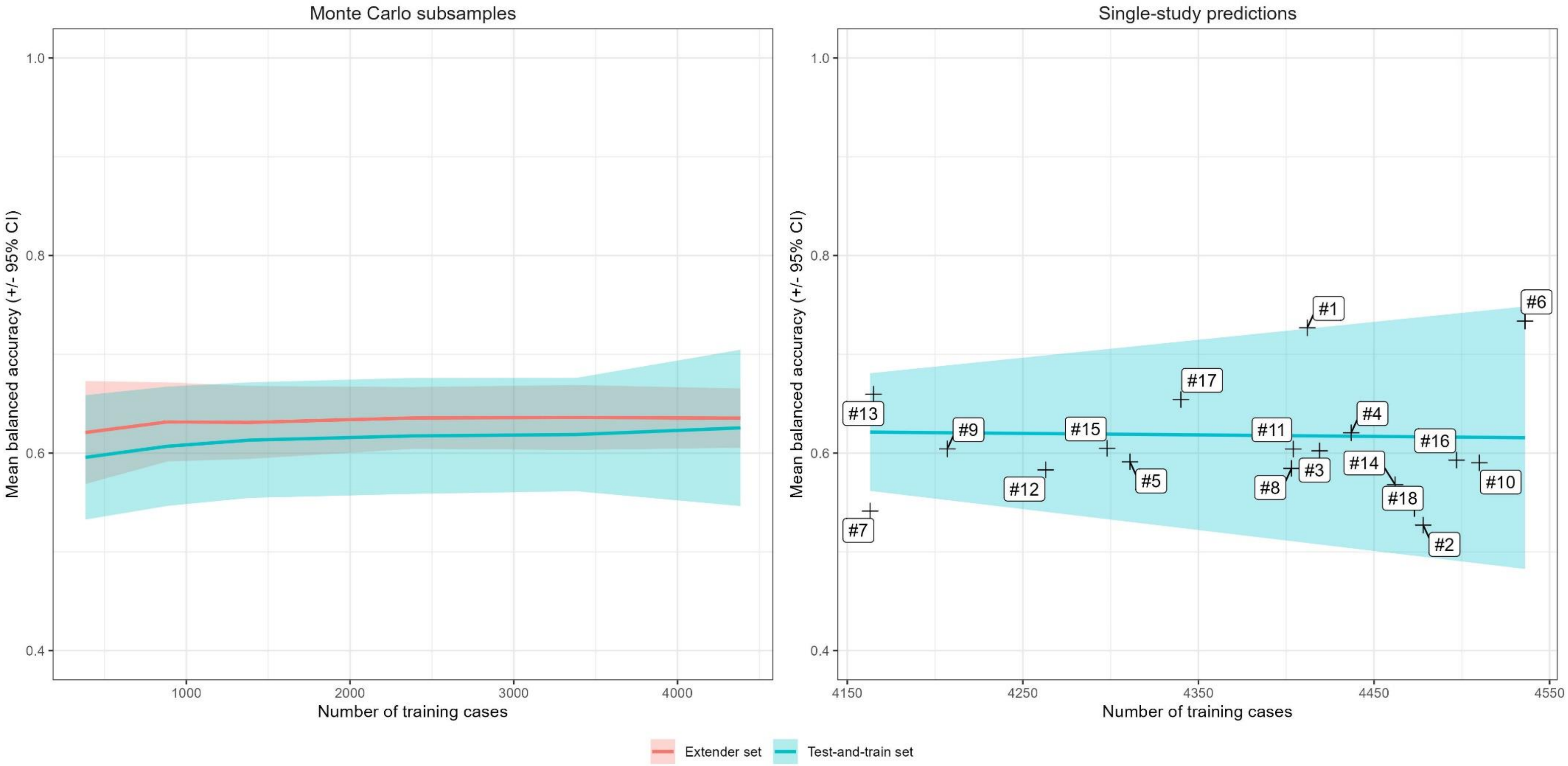


*The left panel gives mean and 95% confidence interval (CI) BAC for subsampled training sets ranging in size from 384 to 4384 cases. The right panel shows the same for subsampled and single-study BAC for leave-one-study-out analyses. Each number #1 to #18 represents one study in the train-and-test set (see Test-and-train set under **Materials and methods**).*

**Supplementary figure 5 – BAC for xgbTree model (subsampled test data, extender test data, and single-study data)**

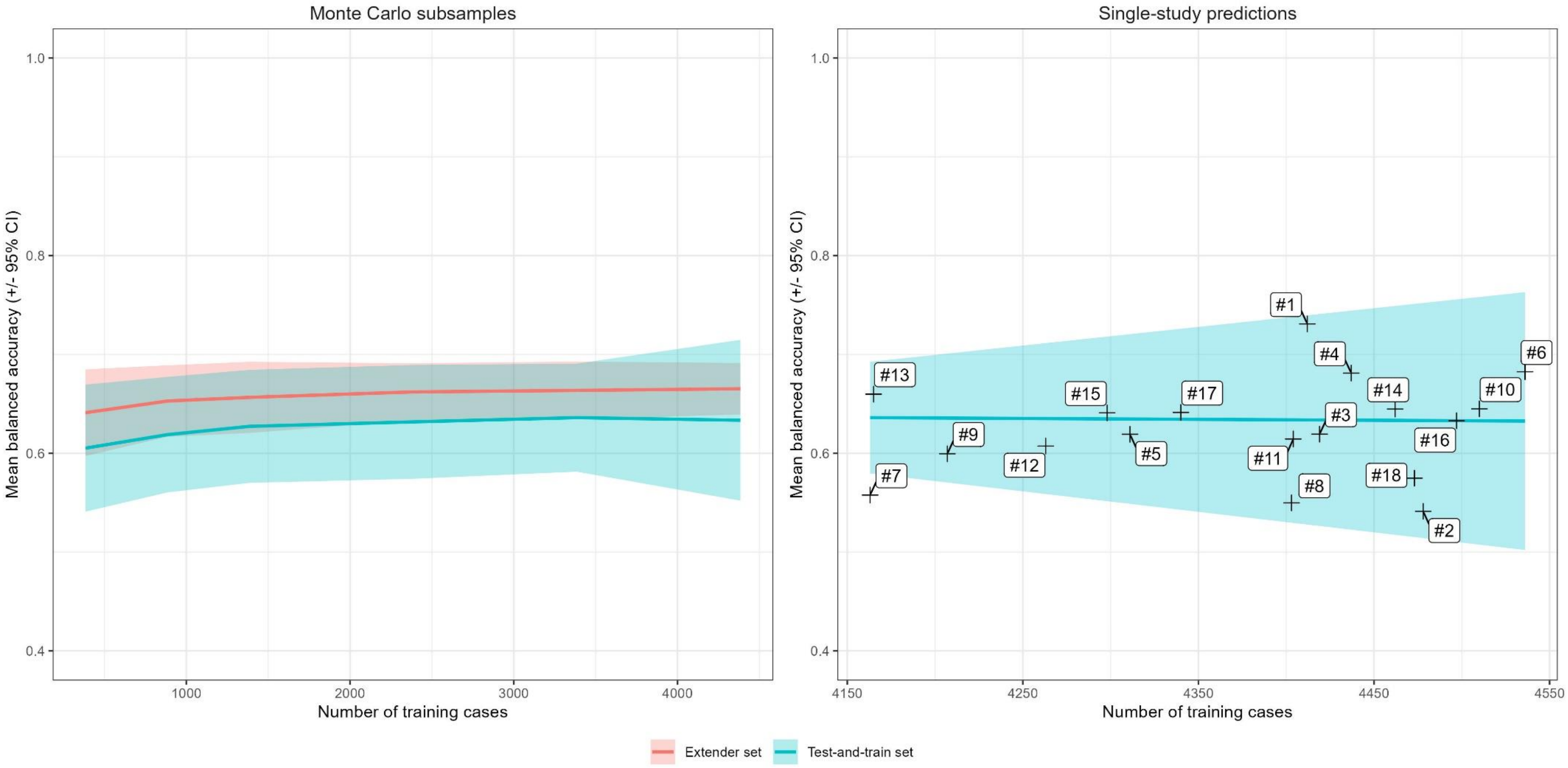


*The left panel gives mean and 95% confidence interval (CI) BAC for subsampled training sets ranging in size from 384 to 4384 cases. The right panel shows the same for subsampled and single-study BAC for leave-one-study-out analyses. Each number #1 to #18 represents one study in the train-and-test set (see Test-and-train set under **Materials and methods**).*

### Additional references


1. United States Food & Drug Administration. Drug Approval Package - Risperdal. 1993. Not currently available online. Available via request to the United States Food & Drug Administration or by contacting the authors.
