## Supplementary figures and images for "The path toward generalizable clinical prediction models"

### Figure 1 (PDF format)

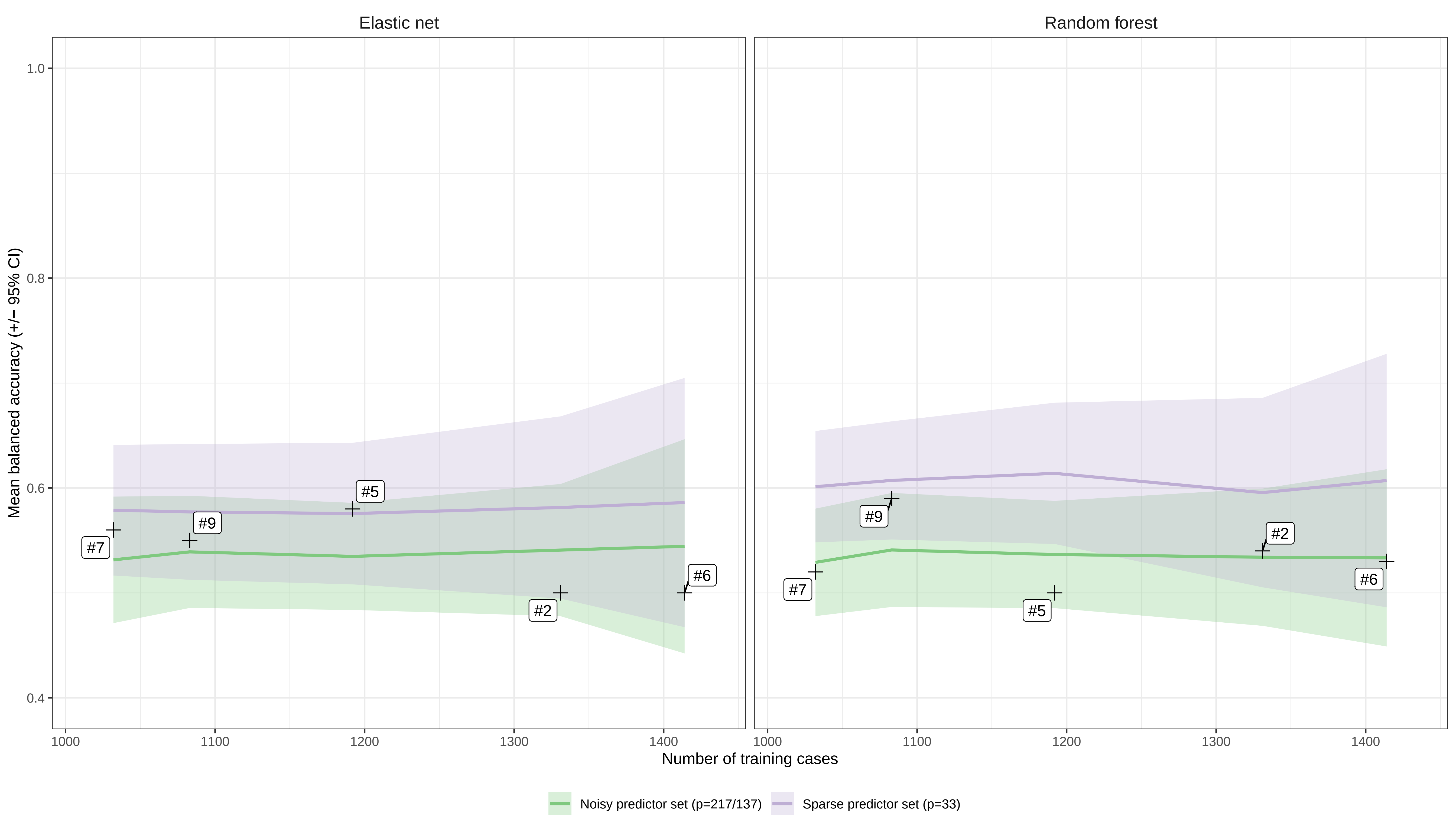

### Figure 2 (PDF format)

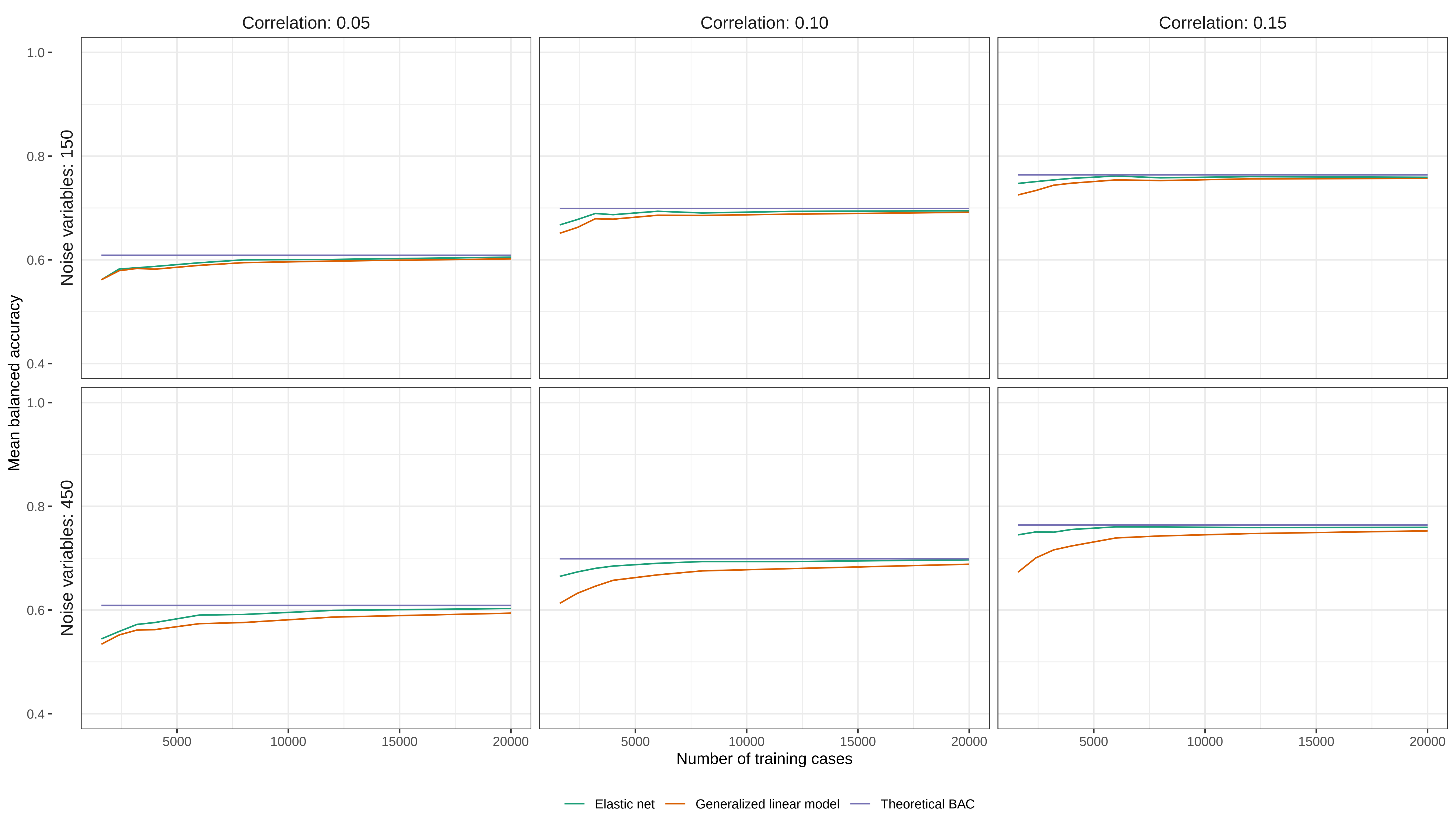

### Figure 3 (PDF format)

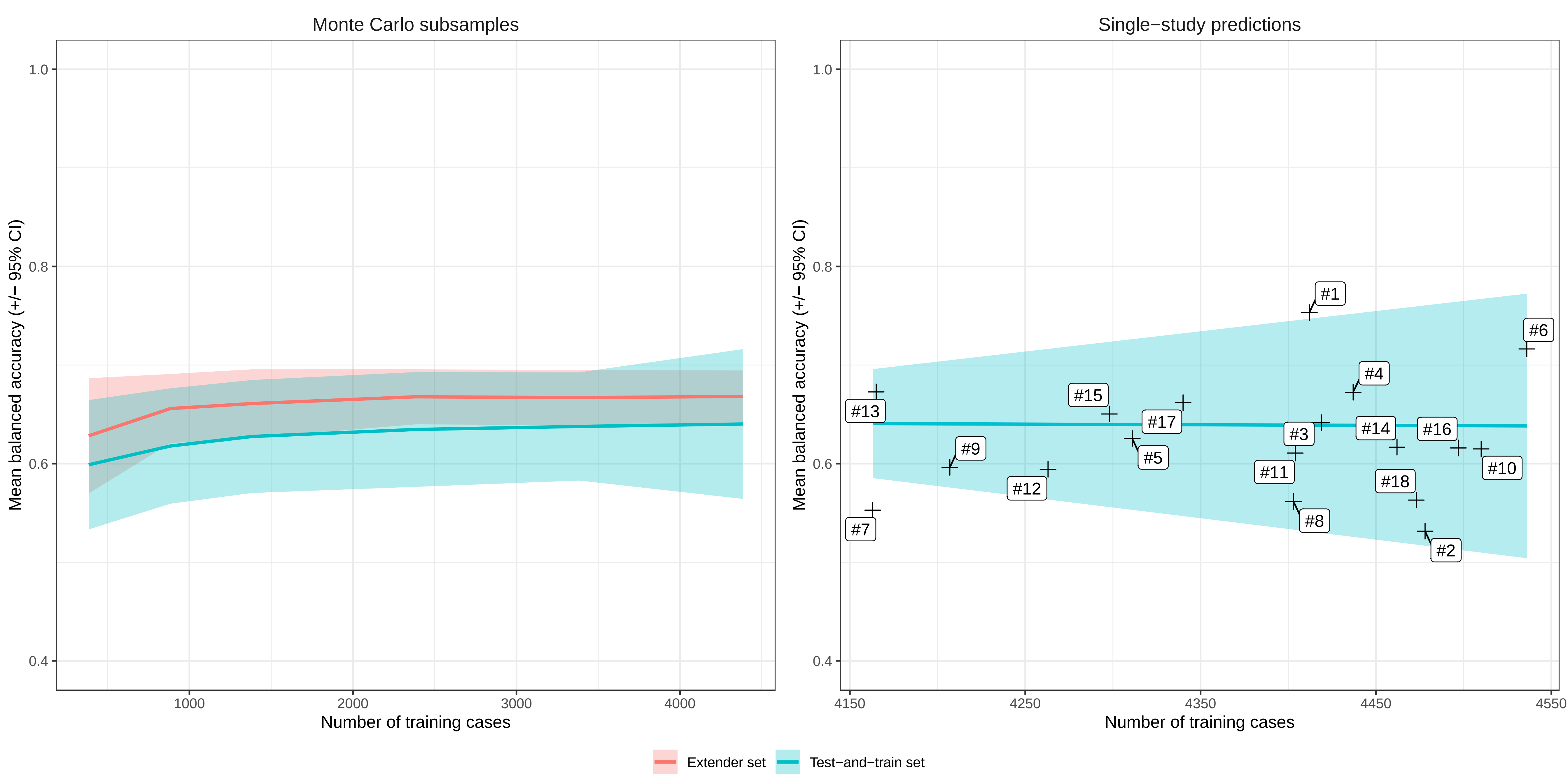

### Supplementary figure 1 (PDF format)

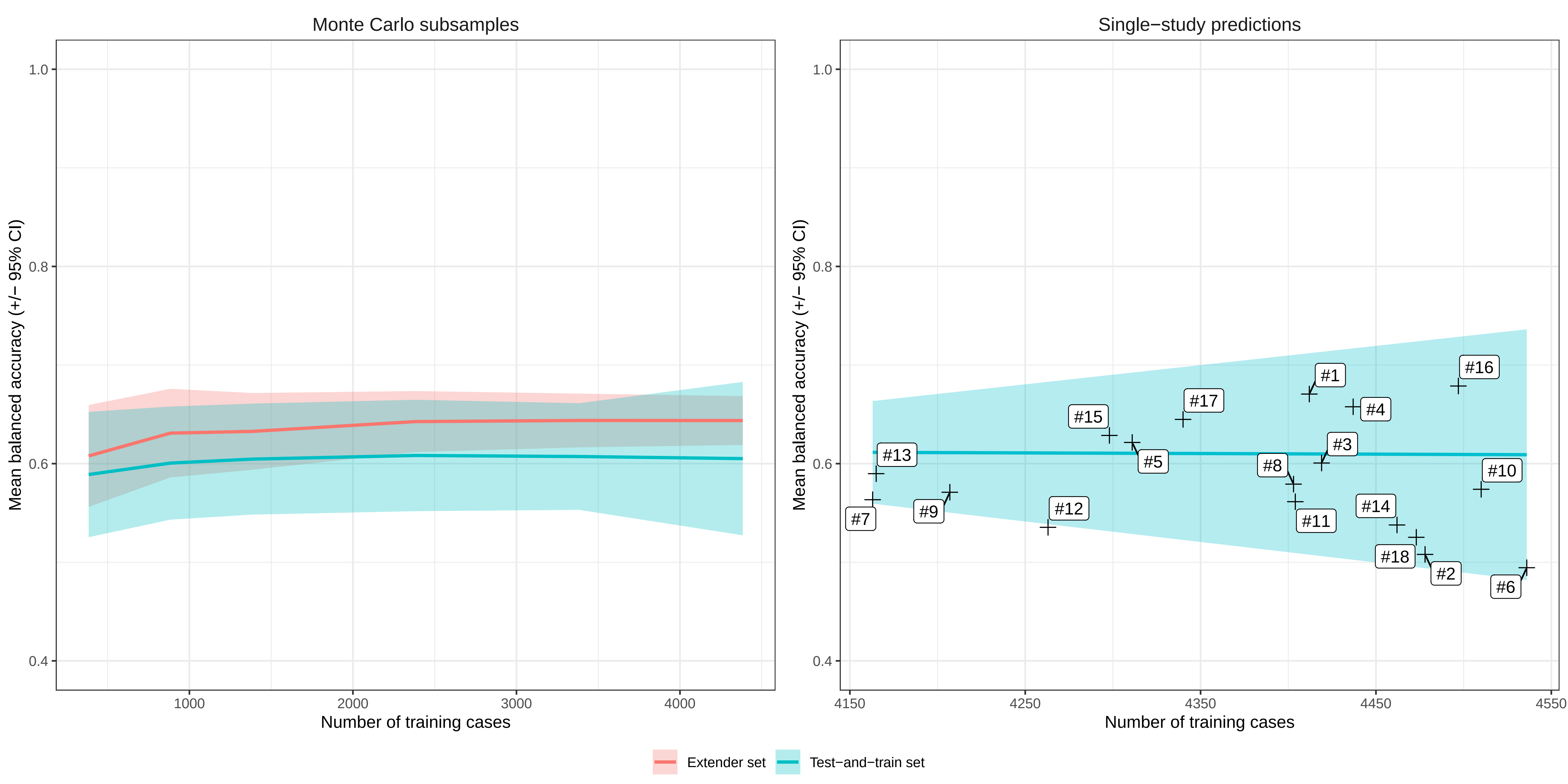

### Supplementary figure 2 (PDF format)

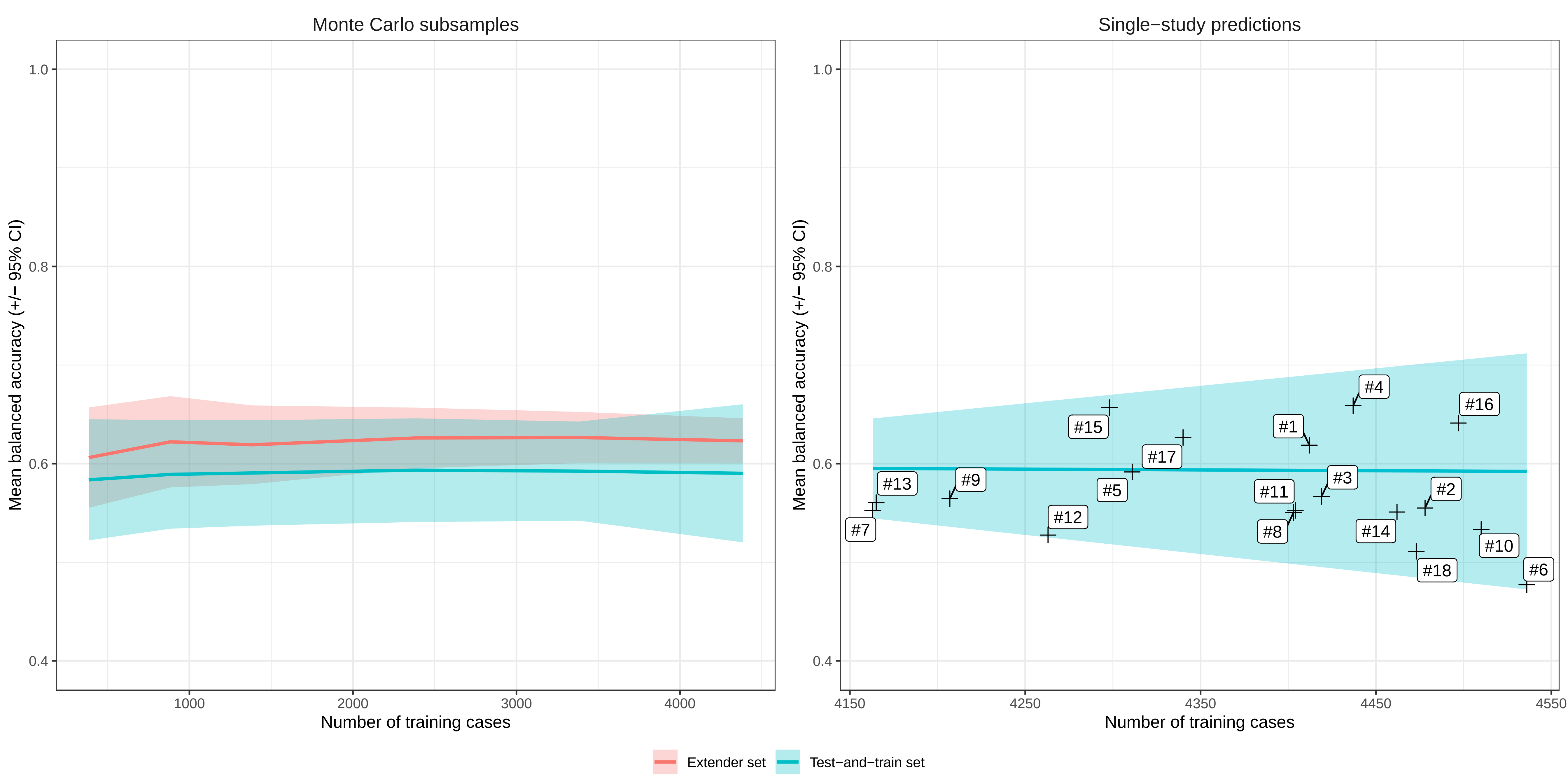

### Supplementary figure 3 (PDF format)

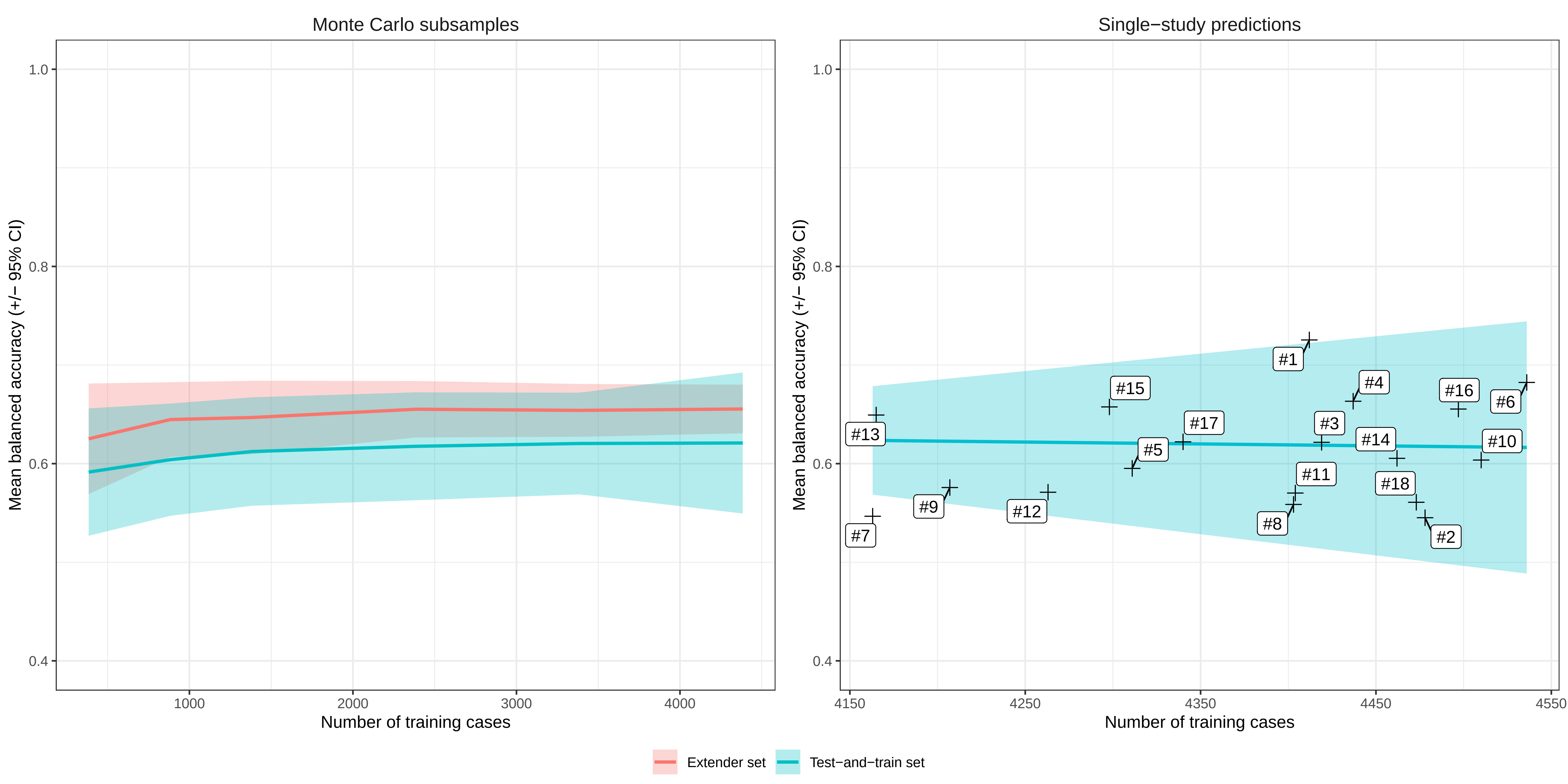

### Supplementary figure 4 (PDF format)

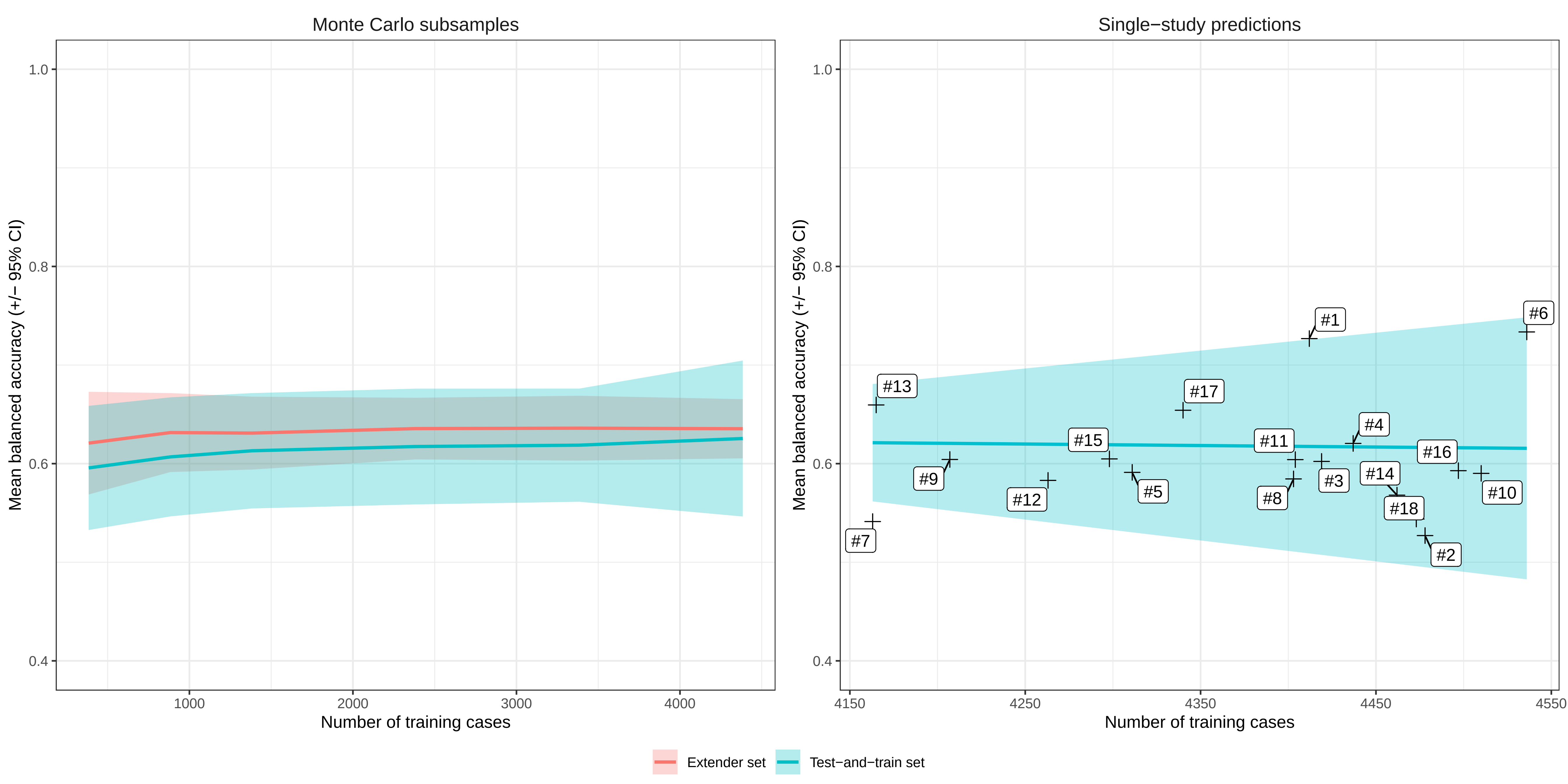

### Supplementary figure 5 (PDF format)

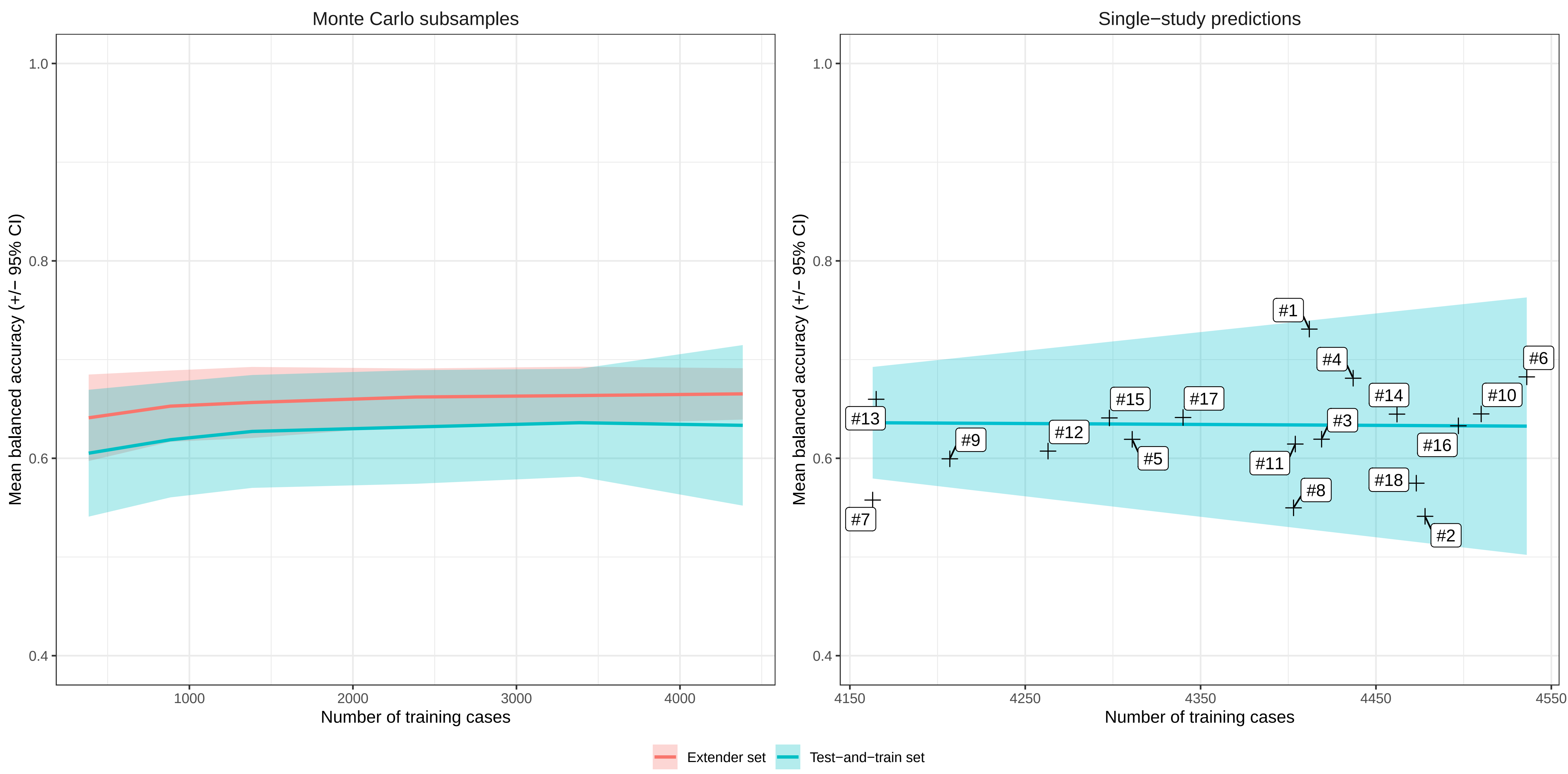
